# Combination of personalized computational modeling and machine-learning for optimization of left ventricular pacing site in cardiac resynchronization therapy

**DOI:** 10.1101/2022.12.14.22283450

**Authors:** Arsenii Dokuchaev, Tatiana Chumarnaya, Anastasia Bazhutina, Svyatoslav Khamzin, Viktoria Lebedeva, Tamara Lyubimtseva, Stepan Zubarev, Dmitry Lebedev, Olga Solovyova

## Abstract

**Background:** The 30-50% non-response rate to cardiac resynchronisation therapy (CRT) calls for improved patient selection and optimized pacing lead placement.

**Objective:** The study aimed to develop a novel technique using patient-specific cardiac models and machine learning (ML) to predict an optimal left ventricular (LV) pacing site (ML-PS) that maximizes the likelihood of LV ejection fraction (LVEF) improvement in a given CRT candidate. To validate the approach, we evaluated whether the distance D_*PS*_ between the clinical LV pacing site (ref-PS) and ML-PS is associated with improved response rate and magnitude.

**Materials and Methods:** We reviewed retrospective data for 57 CRT recipients. A positive response was defined as a more than 10% LVEF improvement. Personalized models of ventricular activation and ECG were created from MRI and CT images. The characteristics of ventricular activation during intrinsic rhythm and biventricular (BiV) pacing with ref-PS were derived from the models and used in combination with clinical data to train supervised ML classifiers. The best logistic regression model classified CRT responders with a high accuracy of 0.77 (ROC AUC=0.84). The LR classifier, model simulations and Bayesian optimization with Gaussian process regression were combined to identify an optimal ML-PS that maximizes the ML-score of CRT response over the LV surface in each patient.

**Results:** The optimal ML-PS improved the ML-score by 17*±*14% over the ref-PS. Twenty percent of the non-responders were reclassified as positive at ML-PS. Selection of positive patients with a max ML-score >0.5 demonstrated an improved clinical response rate. The distance D_*PS*_ was shorter in the responders. The max ML-score and D_*PS*_ were found to be strong predictors of CRT response (ROC AUC=0.85). In the group with max ML-score>0.5 and D_*PS*_ < 30 mm, the response rate was 83% compared to 14% in the rest of the cohort. LVEF improvement in this group was higher than in the other patients (16±8% vs 7±8%).

**Conclusion:** A new technique combining clinical data, personalized heart modelling and supervised ML demonstrates the potential for use in clinical practice to assist in optimizing patient selection and predicting optimal LV pacing lead position in HF candidates for CRT.

## 1 INTRODUCTION

In addition to being at optimal medical treatment, cardiac resynchronization therapy (CRT) is an effective therapy for selected patients with chronic heart failure (HF). In the conventional configuration, CRT delivers biventricular (BiV) pacing to correct electromechanical dyssynchrony of the ventricles in order to increase cardiac output. Despite the well-documented CRT benefits for improving patient outcomes and reducing hospitalizations and mortality in CRT recipients, it still remains ineffective in 30-50% of cases (1). Therefore, stronger predictive indications for patient selection need to be further elucidated and justified (2, 3, 4).

Pacing lead configuration for CRT has been shown to be an essential determinant of patient improvement (3, 5). Several approaches have been proposed to guide left ventricular (LV) lead placement for BiV pacing using new imaging techniques compared to routine fluoroscopy. Pre-operative assessment of myocardial fibrosis and scar area has been proposed to avoid this area when implanting the LV lead (6, 7, 8). Suggestions have been made to individualize LV lead placement using characteristics of ventricular electrical activation dyssynchrony derived from standard 12-lead ECG or from novel electrocardiographic imaging (ECGi), such as QRS duration (QRSd) and QRS area, Q-LV (RV-LV) delay, total biventricular activation, inter-ventricular uncoupling, activation delay vector, and late activation time (LAT) area. However, conflicting data have been obtained on the potential utility of these characteristics for predicting the acute and chronic clinical CRT response (9, 10, 11, 12, 13, 14). This highlights the need for further development and validation of useful multimodality imaging-guided strategies for CRT optimization.

Predictive models have been developed using modern machine learning (ML) approaches to estimate CRT mortality or hospitalization risks from baseline clinical parameters (15, 16, 17), to stratify candidates, and to assess postimplant outcomes (18, 19, 20). Recently, an ML calculator of CRT response based on a minimal set of conventional preoperative clinical data was developed and tested on a large patient population, showing a high accuracy in predicting postimplant improvement in the LV ejection fraction (LVEF) (21).

Modelling studies have used detailed anatomical models of the heart to identify ventricular activation features predictive of CRT improvement (22, 23). A recent article by Rodero et al. (24) proposed a model-based approach to selecting an optimal LV pacing site for a quadripolar LV lead by minimising the total ventricular activation time in personalised cardiac electrophysiology models. Using personalized computational cardiac models and machine-learning (ML) techniques, the RV-LV electrical delay and mechanical regional time to peak contraction were shown as predictors of an acute hemodynamic response to BiV pacing (25).

In all the studies mentioned, the question of pacing configuration is considered independently of and after the patient selection decision. In the current article, we develop a novel technique using a combination of clinical data, computational modelling and ML that may help to solve both problems simultaneously during preoperative patient evaluation: to assess the probability of CRT response for a given patient and to suggest an optimal pacing lead configuration to guide implantation if the patient is selected. We hypothesised that characteristics of ventricular activation during BiV pacing, derived from simulations in validated personalised cardiac models, can be used in combination with baseline clinical data to classify a patient as a potential responder/non-responder to pacing. In parallel, the technique identifies an optimal LV pacing site position for the individual patient that maximises the probability of a positive response.

In our recent paper (26), we developed supervised classifiers of CRT response using ML algorithms, trained on a combination of clinical data and model-derived features (Fig. 1). The best classifier generated an ML-score predicting the probability of CRT response, defined as more than 10% LVEF improvement during chronic BiV pacing, with a high accuracy (ROC AUC=0.82), outperforming classifiers based on pre-implant clinical data alone. As input data, the ML classifier used selected model-derived indices depending on the LV pacing site position, such as the distance from the LV pacing site to the scar area and the RV-LV activation delay during BiV pacing. Since the ML-score is dependent on the LV pacing site, we decided to use it to assess the probability of CRT response at any available position across the LV surface and to find an optimal pacing site that maximises the probability of response.

**Figure 1.**
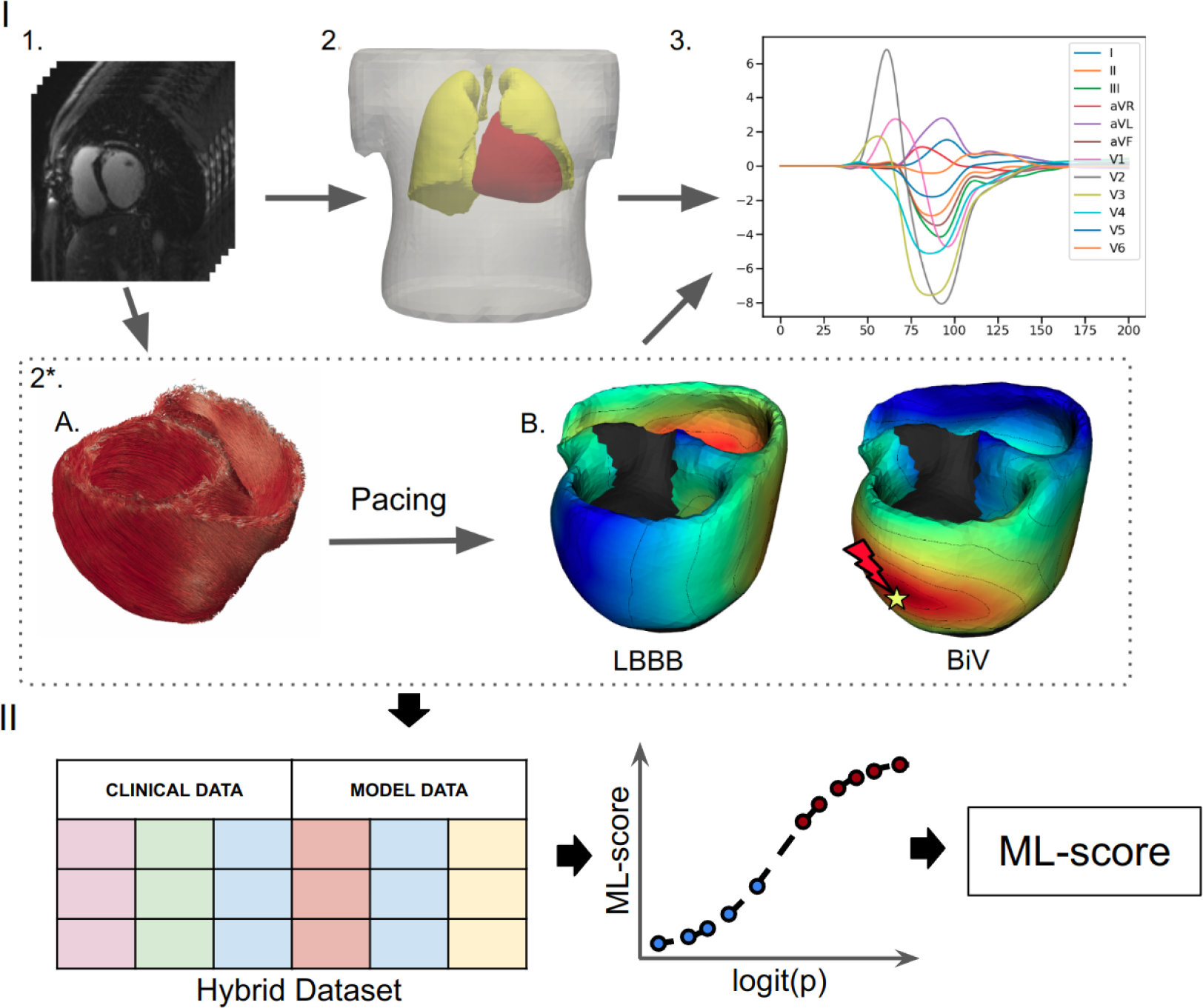
Schematic outline of ML model development. I. Building and calculation of a personalized electrophysiological ventricular model: 1. Processing of the CT imaging data. 2. Segmentation of the finite element meshes of the torso, lungs and ventricles; 2*. Personalization of the ventricular model: A. Rule-based generation of myocardial fibers. B. Assignment of the scar/fibrosis area in the ventricles (shown in back) and computing of the ventricular activation map at the baseline LBBB pattern and BiV pacing with clinical lead position. 3. Calculation of ECG signals from the ventricular activation map. II. Development of a supervised machine learning classifier: creation of a dataset contacting combination of the clinical data and simulated features from the electrophysiological model from each of the 57 patients labeled into responders and non-responders, supervised training of a ML classifier and calculation of the ML-scores of CRT response.

Here, we aimed to further develop a general concept of using a combination of clinical data, patient-specific model simulations and ML techniques to improve patient stratification and guide optimised clinical interventions. The concept is based on the idea of using personalised cardiac mechanistic models for preoperative testing of any clinically relevant conditions of cardiac activity (e.g. excitation patterns, mechanical loading, haemodynamic conditions) or interventions (e.g. pacing, radiofrequency ablation, pharmacology) to predict adverse events or assess the effects of therapy from the simulation results combined with ML predictive models. Comprehensive model testing can reveal a solution that predicts the consequences of interventions, which can aid decision making and/or assist in treatment planning and optimisation. Several exciting examples of the application of this concept were presented by N. Trayanova’s laboratory, which developed a model-based and ML technique for predicting arrhythmia inducibility in post-infarction patients (27), predicting the risk of arrhythmic sudden cardiac death in patients with ischaemic heart disease (28) and cardiac sarcoidosis (29), assessing the likelihood of atrial fibrillation recurrence after pulmonary vein isolation (30), and other clinical applications.

To the best of our knowledge, we are the first to apply such a concept to develop an ML-based technique for CRT optimisation aimed at the preoperative identification of an optimal LV pacing site that predicts the maximum probability of CRT response during BiV pacing. A novel feature of our proposed methodology is the use of a multivariate ML classifier trained on the combination of clinical and model-derived data to predict LVEF improvement in chronic CRT recipients, to optimise the probability of CRT response (ML score generated by the ML prediction model) for a given patient by varying the available LV pacing site location across the LV surface, and to identify and visualize the optimal LV pacing site location. If the maximum ML score predicts a negative response, the patient should probably not be recommended for CRT. Conversely, a patient classified as positive could be proposed as a candidate for CRT implantation and the area of the LV surface with the maximum ML score could be considered as the target for lead implantation. The design of our methodology allows it to be further improved and extended using available and emerging clinical and simulation data, comparing different CRT response criteria and alternative pacing strategies to predict the best way to treat the patient.

To validate our approach, we found a higher rate of the clinical CRT response in patients classified as positive by the maximum ML-score prediction. We also showed that a short distance from the site of the implanted LV pacing lead to the optimal position suggested by our algorithm was a strong predictor of the chronic response in our patient cohort.

Thus, the contribution of our proof-of-concept study is to demonstrate the potential of an ML technique using simulations of personalised computational models to improve patient selection for CRT implantation and to suggest a pacing optimisation strategy in selected candidates.

## 2 METHODS

The pipeline for an ML technique using for BiV pacing optimization is as follows (Fig. 2). A personalized ventricular model for a given patient is constructed using imaging data and is then used to calculate ML-scores generated by a ML classifier of CRT response for various LV pacing sites located on the epicardial surface of different LV segments with the exception of labeled scarring regions. Then Gaussian process regression is applied to the ML-score array to find the locations of pacing sites at the LV surface predicting positive/negative response to BiV pacing and to identify an optimal pacing site that maximizes the ML-score of CRT response for the patient. The maximum ML-score is used to classify the patient as a positive or negative CRT responder. In case of positive prediction, the area of positive ML-scores on the LV surface is visualized with the location of the pacing site with the maximum ML-score indicated. The ML-score map can be used to target LV lead implantation.

**Figure 2.**
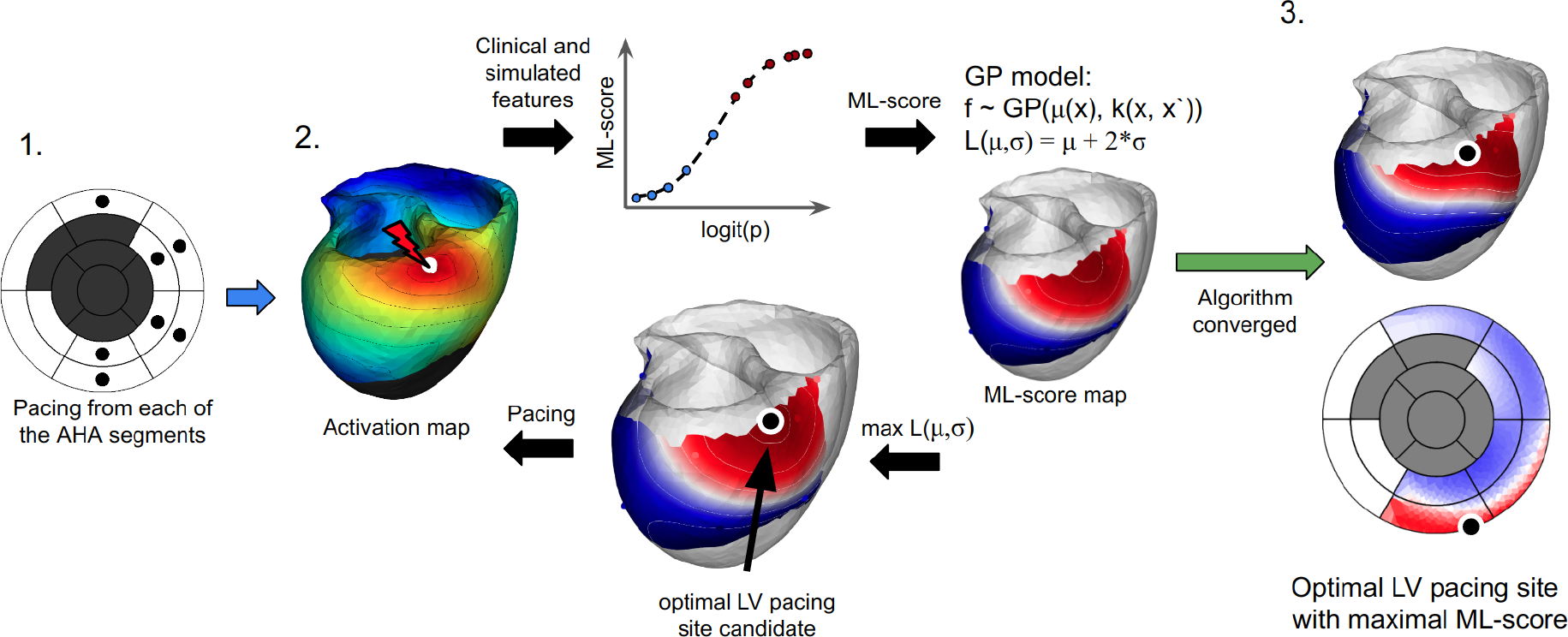
The algorithm for finding the optimal LV pacing site involves three major steps: 1. To compute ventricular activation maps (color map shows activation times of the ventricular regions with early activated regions shown in red and late activated areas shown in blue) from the personalized model at BiV pacing with LV pacing site (white dot) located at every centre of the AHA LV model (excluding the septal and postinfarction scar segments marked as dark gray on the left panel). 2. To apply an LR classifier of CRT response and iterative Bayesian optimization procedure with Gaussian process regression to predict ML-scores on the LV surface. Model-derived features and patient clinical data for each LV pacing site are fed to an LR classifier(shown schematically as a graph in the middle top. The plot shows the LR function calculated for each LV pacing site tested. The x-axis shows a linear combination of the input features (logit(p)) used to calculate the LR value (ML-score). Red and blue dots show positive (red) and negative (blue) predictions of CRT response based on ML-score ><0.51). The LR classifier generates an initial array of ML-scores for interpolation. A Gaussian process regression model is then trained to estimate the GP acquisition function L(mu,sigma) and predict ML-score values on the entire LV surface (see two color maps on the LV surface with shades of red for ML-score>0.5 and shades of blue for ML-score<0.5). The target point candidate is found by approaching the maximum value of the acquisition function (black dot). A new ventricular activation map and simulated features are computed at BiV pacing with the LV site located at the current candidate point. The simulated features in the next iteration step are fed again to the LR classifier to generate an ML-score and retrain the GP regression with accounting for this value or further interpolation of ML-score on the LV surface. 3. The algorithm converges if two iterations predict the same candidate point. The last point with the maximum ML-score value provides an optimal LV pacing site (black dot). The resulting ML-score map is displayed on the LV surface of the personalized LV model and the LV AHA segment scheme.

In what follows, we first describe methods used to develop a ML classifier of CRT response. Next, we describe a technique we developed for LV pacing site optimization. The ML classifier is used as a key tool in the implementation of the CRT optimization technique.

### 2.1 ML classifier of CRT response

In the present study, we used our hybrid data approach to develop a supervised classifier of CRT response as previously described (26) (Fig. 1). The ML classifier was trained and tested on a hybrid dataset consisting of clinical data from CRT recipients and simulated data from personalized computational models of cardiac electrophysiology.

#### 2.1.1 Clinical data

The study involved clinical data from 57 HF patients. The same cohort of patients has been involved previously and described in detail in our recent paper (26), so here we include the summary of the group statistics in the Supplementary material. All patients were on optimal drug treatment following CRT device implantation at Almazov National Medical Research Centre between August 2016 and August 2019. The participants signed approved informed consent forms. The study protocol was approved by the Institutional Ethical Committee. The criteria for patient inclusion in the study and the complete list of clinical data used to perform feature importance analysis for ML classifier development are presented in the Supplementary Material (sec. Clinical data description).

In addition to the standard protocol of patient evaluation for CRT implantation, we also acquired data from 12-lead ECG and echocardiography recordings prior and after device implantation. Computer tomography (CT) was performed to visualize the torso and the heart. The series captured with a scanner (Somatom Definition 128, Siemens Healthcare, Germany) were imported into special Wave program version 2.14 (Amycard, EP Solutions SA) to reconstruct the 3-dimensional geometry of the torso and heart. Finally, epi/endo ventricular surface models were manually built and active pacing sites for the RV and LV leads for BiV pacing were derived from the CT scans and used as reference for model simulations. RV electrodes were placed at a standard apical position in all the patients. Figure S1 (here after prefix S denotes figures and tables in the Supplementary material) shows the distribution of the LV pacing sites between the segments according to the 17 segment AHA LV model. In 50 (88 %) out of the 57 cases, the LV lead was placed in the lateral wall, mostly in the mid- and basal segments. The LV lead was delivered to the inferior segment in only one case, and to the anterior segments in 5 cases. In 2 participants, an apical LV lead position was observed.

Data from magnetic resonance imaging (MAGNETOM Trio A Tim 3 T, Siemens AG or INGENIA 1.5 T, Philips) with contrast (Gadovist or Magnevist) before CRT implantation were used to detect scar/fibrosis areas in the myocardium and to incorporate these data into personalized ventricular myocardial models. Figure S2 shows the distribution of the segments with scar and fibrosis between the 17 AHA LV segments in the patient cohort.

Patients were evaluated before CRT device implantation and during the follow-up period of 12 months after implantation. The clinical data in intrinsic sinus rhythm (baseline) and during BiV pacing in the patient cohort are presented in Table S1 in Supplementary Materials.

##### Responders and non-responders

Patient data were annotated into responder (n=23/40%) and non-responder (n=34/60%) groups according to LVEF improvement by more than 10%. Table S2 shows clinical data in the groups, indicating significant differences in the echocardiography indices.

#### 2.1.2 Simulated data

##### Ventricular anatomy models

Based on the segmentation of CT imaging data, finite element models were constructed for the torso, lungs and RV-LV ventricles for each of the 57 patients (Fig. 1, panels I.1-2) . A rule-based approach was used to simulate myocardial fiber architecture (31). MRI data on scarring and fibrosis areas in the myocardium were accounted for in the LV model using expert annotation of these areas within the 17-segment American Heart Association (AHA) model of LV (see (26) for more detail). Similar to many other modelling studies (e.g.(32, 33, 34)), the scar regions were simulated as non-conducting and non-excitable areas, and the conductivity of the fibrosis regions was decreased by 50%. Limitations of these assumptions and further directions for model improvement are discussed in Section 5 below.

##### Models of myocardial electrical activation and ECG

Like in the previous work (26), we used an Eikonal model (35) to calculate electrical activation times at each point on the ventricular mesh. Cardiac tissue was simulated as an anisotropic medium with conductivities resulting in an excitation velocity ratio of 4:1 along vs across the myocardial fibers. The Eikonal model is currently widely used; it allows one to simulate the evolution of the cardiac excitation wavefront (36, 37, 38, 39, 40). To compute ECG signals, we calculated a map of potentials in the heart, by assigning a predefined cellular action potential to each model element at corresponding activation time. The widely used human ventricular action potential model TP06 (41) was taken as a model of cardiomyocyte action potential generation. The TP06 provides a mathematical description of the main ionic currents that are involved in AP genesis and has been thoroughly validated in the literature. ECG calculation was performed using the Lead Field method proposed by Pezzuto et al. (37, 40). ECG signals were computed according to the standard 12-lead ECG definition and the lead-field approach allowed us to reduce calculation time more than 100x times.

##### Pacing protocols

We simulated two pacing protocols – baseline activation pattern at LBBB and BiV pacing. The model-derived features of the ventricular electrical activation were then used for developing an ML classifier of CRT response and for searching for an optimal LV lead location.

To simulate the LBBB activation pattern, the RV endocardial surface was annotated and a Purkinje network with excluding left bundle branch was generated using the model proposed by Costabal et al. (42). Note, that in this study we assumed the total LBBB for all patients and thus excluded the left branch from the Purkinje network in the models. The Purkinje system was isolated from the working myocardium and connected to it only at the ends of the Purkinje fibers. Activation started at the His node and spread throughout the conduction system with an excitation velocity of 3 mm/ms before approaching the Purkinje-myocardial junction points. This activation map was then applied to initiate activation within the ventricular myocardium according to the Eikonal model. Furthermore, we used the simulated LBBB ventricular activation map to define the area of late activation time (LAT) in every patient model. The distance from the LAT area to LV pacing site was determined and tested as an optional measure to predict the optimal LV lead location (see next section below).

For simulations of BiV pacing with no interventricular delay, we used the locations of RV and LV pacing sites, manually segmented from the CT images.

For both the LBBB and BiV protocols, clinically recorded maximum QRSd were utilized to personalize the global myocardial conductivity parameter in each model. In this study, we used this rather quick to implement but representative approach to fit simulations to physiological data. Here, we did not use more detailed data on QRS morphology or other ECG features to identify the electrophysiological parameters in our personalised models. Our study design was focused on predicting the effects of BiV pacing on several integrative features of the ventricular activation and its dyssynchrony, reflected in the change in QRSd, which was accurately fitted in the models. We employed the L-BFGS-B algorithm to handle optimization in the model and the method proposed in (43) for automatic QRS onset and offset detection.

##### Consistency of the ventricular activation simulations with clinical data

The personalized ventricular models yielded average 12-lead QRSd values very close to the clinical data (correlations between simulated and clinical QRSd: r=0.99 at baseline and r=0.99 at BiV pacing, p<0.01, see Fig.S6,S7 in the Supplementary materials). Furthermore, the model simulations showed a congruence with the clinical QRS morphology and realistic qualitative patterns of ventricular activation with the typical U-shaped activation at baseline LBBB and realistic patterns and synchronisation of activation during BiV pacing (see Fig. S8 in Supplementary materials). To validate the electrophysiology model predictions we also used data from non-invasive ventricular mapping (ECGi, Amicard 01, EPI-solutions) performed for several patients from our cohort. In a majority of models (15 (75%) out of 20 models with LAT segments available from ECGi), LAT area at baseline LBBB defined from simulated activation map was concordant or adjacent with that defined from ECGi. In addition, our models yielded total activation time of the ventricles that correlated with that predicted by ECGi (r=0.96 for LBBB, r=0.80 for BiV, p<0.001, see Fig.S9,S10). The personalized models also clearly captured clinically seen synchronisation of ventricular activation during BiV pacing as revealed in the reduction in the individual and average simulated indexes of the ventricular electrical dyssynchrony: total activation time, inter- and intra-ventricular activation delay (see TAT_95_, AD*_RV LV_*, and AD*_ST LV_* in Tables 1 and S1). The consistency of the model simulations with the clinical data at baseline LBBB and during postoperative BiV pacing suggested the possibility of using model-derived indices in combination with clinical measures to develop ML classifiers for predicting CRT response.

##### Simulated features used for developing ML classifiers

The following model-derived indices were used as measures in CRT response prediction (see Tables S1, S2 in Supplementary Materials). The first group of model-derived indices was derived from the ventricular anatomy models based on CT and MRI data, coupled with electrophysiological model simulations. We estimated the volume of postinfarction scar and non-ischemic fibrosis and their size relative to the myocardial tissue volume. Knowing the exact locations of the RV and LV pacing leads, we computed the time delay in the activation of the LV electrode later than the RV electrode (RV-LV delay) in baseline LBBB. We also calculated the spatial distances between the RV and LV pacing sites (RV-LV distance), and the distances from the LV pacing site to the scarring area (Scar-LVPS distance) and to the area of LAT (LAT-LVPS distance), by solving an isotropic Eikonal equation.

The second group of model-derived indices were derived from simulated activation maps and 12-lead ECG signals in baseline LBBB and during BiV pacing. The following ventricular activation characteristics were considered: maximum QRSd; total activation time of 95% myocardial tissue volume for the biventricular model (TAT95); RV-LV activation delay (AD*_RV LV_*) as the difference between LV and RV total activation time characterizing inter-ventricular electrical uncoupling; intra-ventricular dyssynchrony index as the relative difference between the mean activation time of the LV free wall and the septum (AD*_ST LV_* = (mean AT*_LV lat_* - mean AT*_ST_*)/TAT). Changes in the indices under BiV pacing in comparison with baseline (delta), in either absolute values or normalized to the baseline were also used for developing the ML classifier of CRT response.

#### 2.1.3 Machine learning model

In the previous paper (26), we performed a comparative analysis of several ML supervised classification algorithms applied to our relatively small dataset. Based on the analysis, we chose here a logistic regression (LR) classifier as the easiest to interpret and most robust in terms of overtraining, while showing a high performance similar to more complex ML models.

The LR classifiers were trained on a hybrid dataset containing the clinical and model-derived indices described above. At the preprocessing step, non-categorical data were normalized by substracting the mean and dividing by standard deviation. Highly correlated features were also removed from the dataset by a threshold > 0.85.

To train the LR models, the dataset was labeled into responders and non-responders according to the chronic clinical CRT response defined from the post-operative data as an increase of more than 10% in LVEF (26). We developed the LR classifier using Leave-One-Out cross-validation and three different feature selection methods inside the cross-validation loop to train the classifier. The full list of clinical and simulated features fed to the ML algorithms is shown in Figure S3, sorted by automated feature importance scoring. Using the Leave-One-Out cross-validation approach, we trained multiple LR classifiers on the training datasets each containing all records from the full dataset, but excluding one record for a given patient. The excluded patient features were used to test the corresponding LR classifier and predict the ML-score for that patient. Such LR classifiers were developed for each leave-one-out dataset and tested on each patient from the cohort to estimate the feature importance and accuracy of the final LR classifier. We used feature selection within the cross-validation loop to eliminate any bias factors. In addition, in this study we used a simple LR model that does not tend to overfit on small datasets. Also, we didn’t do any hyperparameter search, as a result of which the models could be overfit. Moreover, during the development of CRT response classifiers in our previous work (26), we also tested standard 5-fold cross-validation with 1000 iterations, which showed classifier performance similar to leave-one-out cross-validation with almost equal ROC AUC and no overfitting.

From the total set of input indices considered for classification, seven most significant features with the highest LR weights were selected as follows. The three pre-operational clinical features were: LVEF (%), body mass index (BMI, dimensionless), and LV end-diastolic diameter (EDD, mm). The four model-derived features were: distance from LV pacing site to scarring area (Scar-LVPS distance, mm), total biventricular activation time (TAT95, ms), and RV-LV activation delay (AD*_RV LV_*, ms) at LBBB and during BiV pacing. Note, that two out of the four model-derived indices, Scar-LVPS distance and AD*_RV LV_* at BiV pacing, depend on the LV pacing site position, and may change with LV pacing site moving on the LV surface. These seven features were used to train the final LR classification model (see Table S3). The LR classifier generates an ML-score that provides an estimate of the probability of a positive CRT response for the patient.

The LR model accuracy was estimated using the area under the receiver operating characteristic curve based on the results of Leave-One-Out cross-validation (ROC AUC, Fig. S3 in Supplementary Materials). A cut-off ML-score of 0.5, which maximizes the accuracy of the LR model, was applied to predict either a positive or negative response to CRT in our patient cohort. This LR classifier was then used for LV pacing site optimization.

### 2.2 Optimization of LV pacing site position based on the ML-score

We used our LR classifier to optimize the LV pacing site position during BiV pacing for each personalised ventricular model. In the current study, we focused on optimising conventional BiV pacing according to the clinical data in patients we used. Therefore, we did not simulate other pacing configurations potentially effective for CRT, such as His-Purkinje conduction system pacing or multiple site pacing, etc., as no clinical data were available to validate such predictions. In our simulations, the RV pacing site was set to a reference position manually segmented from the CT images, as its position did not vary significantly between patients and was typically located in the conventional RV apical region. The position of the LV pacing site was varied over the entire LV epicardial surface available for BiV pacing. Septal regions (as not available for conventional transvenous or epicardial access) and scarred areas (as ineffective) were excluded from consideration. A problem of ML-score optimization across the LV surface was solved. Figure 2 shows the pipeline employed for finding an optimal LV lead position that would maximize the ML-score of CRT response.

First, we varied the position of the LV pacing site between the centers of LV AHA segments on the epicardial surface (Fig. 2, step 1). For each LV lead position (up to 12 positions, 10 per model on average), we computed the personalized electrophysiological model during BiV pacing and extracted model-derived features from the simulations. The pacing site dependent indices along with other input features were fed into the LR classifier to generate the ML-score. At the end of this step, an initial set of ML-score values was collected, characterizing the distribution of the ML-scores in the AHA LV model.

The small number of points with computed ML-scores did not allow us to accurately predict the optimal LV pacing site with maximum ML-score at the LV surface. Therefore, we used a Bayesian optimization method to predict the ML-score values over the entire LV surface accessible for pacing. This method involves building a regression model and its iterative refinement before converging at the optimal solution (44, 45).

#### Bayesian Optimization

The iterative process of the optimal ML-score prediction was performed using Bayesian Optimization with Gaussian process regression (GP regression) model (46). We used the current ML-score set (an initial pre-calculated ML-score vector in the first iteration step) to train GP regression and to predict the ML-score at every node of the mesh on the LV epicardial surface. Then we calculated the so called acquisition function: L(*µ*, *σ*) = *µ* + 2*σ*, where *µ* is an expected ML-score value predicted by GP regression and *σ* is the standard deviation of GP at this point (GP uncertainty value) (Fig. 2, step 2). After that, maximum L(*µ*, *σ*) was found throughout the LV nodes. This LV pacing site was further used to calculate the electrophysiological model at BiV pacing. These model-derived features were fed to the LR classifier to compute the corresponding ML-score.

The Bayesian optimization method thus strikes a balance between finding points that allow one to refine the GP regression model (points with large uncertainty, i.e., large *σ*), and finding points where the value of the regression function is maximum (points with maximum *µ*).

In the next iteration step, GP regression was re-trained with the addition of the ML-score from the new point on the LV surface and the algorithm was repeated. The optimal solution was considered to be found if the last two iterations of the Bayesian optimization algorithm predicted the same point.

Thus, we predict ML-score values across the LR surface grid without the need to compute the model in each grid node (which is time-consuming), and use Bayesian optimisation to iteratively improve a position of the LV pacing site that provides the maximum ML-score across the surface using the GP apparatus and regression model uncertainty information. Note that each iteration step of the algorithm required an additional model calculation at the single point of the current extreme candidate, and in our case the method converged in about 5 iteration steps. Therefore, we consider this approach as reasonable because it is fast, does not require grid refinement, and not only identifies an LV location with maximum ML score, but also predicts all other areas on the LV surface with ML score >0.5 (Ml-score map) that can be considered for LV pacing as positive for CRT response.

Finally, we obtained an LV epicardial surface map of ML-score values (Fig. 2, step 3), predicting areas of LV pacing with either positive (ML-score > 0.5) or negative (ML-score < 0.5) expectation of CRT response and suggested the optimal position of LV pacing site maximizing the ML-score among all available LV surface positions. This map can be used to guide LV lead placement during CRT implantation if the patient is predicted to be a potential responder according to the maximum ML-score > 0.5 and ultimately selected for CRT procedure.

### 2.3 Alternative LV pacing sites

In addition to the ML-score based optimization of LV pacing lead position in the personalized models, we also used an alternative LAT area identified in LBBB for LV pacing site location, as suggested in several clinical studies (47, 48). Another alternative approach to pace our models was based on TAT95 minimization, which was frequently considered as a potential target for LV lead positioning (49). The latter approach was implemented in our personalized models using a similar iterative procedure suggested for ML-score optimization. To this end, we generated an initial set of simulated TAT95 during BiV pacing with LV pacing from the centers of LV segments and then used Bayesian optimization of TAT95 over the available LV surface. As a result, we found an LV pacing site position with a minimal TAT95 for each personalized model of our cohort. The effects of pacing from alternative LV pacing sites were compared with the results obtained for the clinical and ML-based optimal LV lead position.

### 2.4 Software

Cardiac electrophysiology was simulated using an in-house software based on the FENICS library (for solving PDE problems) (50) and VTK (for working with meshes). The scikit-learn library was employed for the machine learning: classifier development, statistical modelling, feature selection, cross validation, and ROC-AUC calculation, and the Pyro (51) library was used for GP regression and Bayesian optimization.

### 2.5 Statistics

Detailed analysis was performed using the IBM SPSS Statistics 23.0.0.0 software package (USA). For qualitative data, the frequency and percentage of total patients in the cohort were calculated. Quantitative data are median [25th-75th] quartiles or mean *±* standard deviation if the criteria for a normal distribution are met. Comparisons between two dependent groups for quantitative data were made using the paired sample t-test for normal distribution and the Wilcoxon test for non-normal distribution and McNemar’s test for qualitative data. Comparisons between dependent groups were made using nonparametric Friedman’s two-way ANOVA, followed by a pairwise comparison adjusted for multiple comparisons. Comparison between two independent groups (non-responders vs responders) was carried out using Mann-Whitney test for quantitative data and Pearson’s chi-square test for qualitative data. Feature dependence was assessed using the Pearson correlation test for normal distribution and the Spearman rank correlation test for non-normal distribution. The critical level of statistical significance was taken equal to 0.05.

## 3 RESULTS

### 3.1 Study population: clinical data and model simulations

We used retrospective data for fifty seven (57) CRT recipients. Clinical follow-up and echocardiographic evaluation were undertaken in one year after implant. The same patient cohort was involved and described in detail in our previous study (26), which focused on the development of ML classifiers of CRT response. Here, the clinical data description, subject characteristics, CT/MRI derived data and model-driven indices in the total patient cohort are presented in the Supplementary Material, Table S1. A summary of the statistics in patients classified into responder or non-responder groups, defined by more or less than 10% improvement in LVEF during follow-up, is shown in Table S2 in the Supplementary material.

Overall, LVEF increased from 26 *±* 6% at baseline to 35 *±* 8 % (paired LVEF improvement (ΔLVEF) of 9*±*8%, p<0.001) with 23 (40%) of the patients classified as responders defined by more than 10% ΔLVEF during follow-up. In the responder group, LVEF increased from 23 *±* 5% at baseline to 40 *±* 6% at followup (paired improvement of 16[14;19]%, p<0.001). In contrast, LVEF improved much less in the non-responder group from 29 *±* 6% to 32 *±* 7% (paired improvement of 4[1;8]%, p=0.002). The only two clinical characteristics distinguished between responders and non-responders at baseline: lower body mass index (27*±*5 versus 30*±*5, p<0.05) and lower LVEF (23 *±* 5% versus 29 *±* 6%, p<0,01) were found in responders compared to non-responders.

A personalized biventricular electrophysiology model accounting for the scarring segments was built for every patient. The clinical (referent) RV-LV pacing sites (ref-PS) were used to simulate BiV pacing in the personalized models. In two (9%) responders and in eight (24%) non-responders the LV segments with implanted electrodes were concordant with scar area. A larger distance from the LV pacing site to the scar/fibrosis zone was revealed in the responders compared with non-responders (44[19;53] mm *vs* 25[3;43] mm, p<0.01, respectively, see Scar-LVPS distance in Table 1). In consistency with the clinical data, a reduction in the simulated QRSd, total ventricular activation time (TAT95, defined as the time of activation of 95% of the ventricular myocardium) and in all computed indices of inter- and intraventricular electrical dyssynchrony was found in the models at BiV pacing against baseline (Table 1, see also Table S2 in the Supplementary material). None of the simulation-based indices of the ventricular activation showed a significant difference between the responders and non-responders at baseline and during BiV pacing.

**Table 1.** Statistics of simulated total activation time 95% (TAT95), QRSd, the ML-score and distance from LV pacing site to the scar area at baseline LBBB activation and BiV pacing at ref-PS and ML-PS in the total patient cohort and groups of responders and non-responders.

| Simulated feature |  | Total Cohort (n=57) | Responders (n=23) | Non-responders (n=34) |
| --- | --- | --- | --- | --- |
| TAT95, ms | LBBB | 151[137;166] | 153[141;164] | 150[133;169] |
|  | ref-PS | 99[88;111]* | 96[88;106]* | 102[88;112]* |
|  | ML-PS | 100[89;114]* | 99[89;113]* | 102[86;114]* |
| QRSd, ms | LBBB | 190[175;205] | 195[180;204] | 189[173;210] |
|  | ref-PS | 142[132;155]* | 142[132;151]* | 147[131;156]* |
|  | ML-PS | 144[132;154]* | 142[136;152]* | 144[129;157]* |
| ML-score | ref-PS | 0.31[0.14;0.68] | 0.73[0.36;0.95] | 0.19[0.07;0.39]<sup>#</sup> |
| | ML-PS | 0.56[0.33;0.87]<sup>\$</sup> | 0.82[0.64;0.98]<sup>\$</sup> | 0.42[0.20;0.73]<sup>,\$</sup> |
| Scar-LVPS distance, mm | ref-PS | 34[10;49] | 44[19;53] | 25[3;43]<sup>#</sup> |
| | ML-PS | 58[45;71]<sup>\$</sup> | 61[51;66]<sup>\$</sup> | 55[41;72]<sup>\$</sup> |
| D<sub>PS</sub>, mm | ML-PS | 50[26;78] | 35[21;61] | 64[41;92]<sup>#</sup> |

Median [25th percentile;75th percentile]
\* - $p < 0.05$ , \* - $p < 0.01$ ML-PS vs LBBB. \$ - $p < 0.05$ , \$ - $p < 0.01$ ML-PS vs ref-PS. Comparison of dependent groups was performed using Friedman's test, followed by a pairwise comparison adjusted for multiple comparisons.

### 3.2 ML classifier of CRT response built on a hybrid dataset

Using a combination of the pre-implant clinical data and indices derived from personalized models of ventricular activation at the LBBB and BiV pacing we built a supervised LR classifier of CRT response (Figure S3 and Table S3 in the Supplementary material). Seven most important features were selected for the final LR classifier. There were three clinical features at baseline: LVEF, BMI, EDD; and four model-derived features: Scar-LVPS distance, TAT95*_LBBB_*, and AD*_RV LV_* at both LBBB and BiV pacing. Two model-derived indices, Scar-LVPS distance and AD*_RV LV_* at BiV pacing depend on the LV pacing site. The LR classifier features a high ROC AUC of 0.84 with a total accuracy of 77%, sensitivity of 65% and specificity of 85% (Table S3). The LR model generates an ML-score as a measure of the probability of CRT response in a patient. The cut-off value of ML-score=0.51 classified patients as responders or non-responders with maximum accuracy.

The ML-score in the total cohort was 0.31[0.14;0.68], with higher values in the responder compared with non-responder group (0.73[0.36;0.95] *vs* 0.19[0.07;0.39], p*<*0.001, Table 1). Moreover, the ML-score values correlate with post-implant improvement in the LVEF (r=0.48, p*<*0.001).

### 3.3 ML-based technique for patient selection and LV pacing site optimization

Utilizing the LR classifier, a novel technique has been developed and implemented to identify an optimal LV pacing site position, which maximizes the ML-score of CRT response for a given patient. The technique combines the following steps (Fig. 2):

1. Compute a patient personalized model at LBBB and during BiV pacing with multiple LV pacing sites located at the centers of the AHA LV model segments, excepting septal segments as unavailable for the conventional transvenous approach and scarring regions as being non-excitable.
2. Extract model-derived features from the simulations depending on the LV pacing site location.
3. Calculate an initial set of ML-scores from the LR classifier fed with input data depending on the pacing site location.
4. Interpolate the ML-score values on the LV surface available for pacing using Gaussian process regression and Bayesian optimization.
5. Find the maximum ML-score and corresponding optimal LV pacing site.
6. Classify the maximal ML-score value into positive (ML-score > 0.5) or negative (ML-score <0.5) for CRT response. Suggest selecting the patient as a CRT candidate according to the model prediction.
7. In case the maximum ML-score is positive, visualize the LV surface map of the ML-score with the labeling of areas of pacing site location predictive of the positive response and indicating an optimal area for LV pacing site.

Figure 3 shows three examples of an ML based optimal LV pacing site (ML-PS) in personalized ventricular models. In each case, a two-color map of the ML-score value is shown on the LV epicardial surface of the personalized biventricular model and on the LV AHA segment scheme. Red shades indicate ML-scores>0.5 on the LV surface predicting a positive response to CRT. In contrast, shades of blue label ML-scores<0.5 unwanted for LV pacing. Blue and red dots on the map show the locations of the clinical ref-PS and optimal ML-PS, respectively.

**Figure 3.**
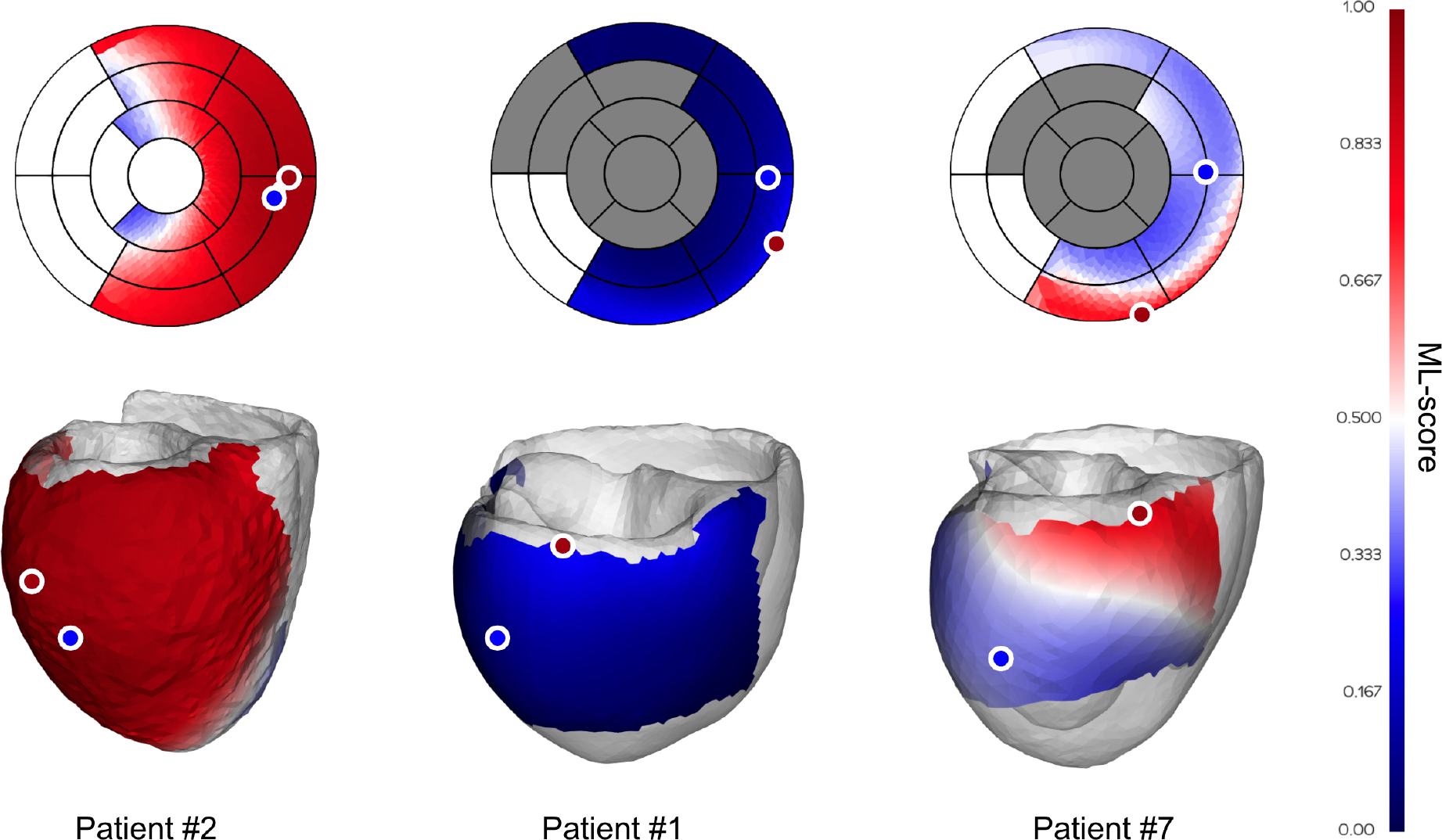
Examples of ML-score map with optimal LV pacing site location in personalized ventricular models. Two color maps of the ML-value are shown on the LV surface of personalized models and on the LV AHA segment schemes. Dark gray at the AHA LV scheme marks segments containing postinfarction scar, which are excluded from pacing. Shades of red show LV surface area with ML-scores>0.5 and shades of blue show ML-scores<0.5. Blue and red dots show the locations of the clinical and optimal LV pacing sites. From left to right are shown examples of the ML-score map in a clinical responder (patient #2), non-responder (patient #1), and non-responder (true negative at the ref-PS) predicted as positive to CRT response at the optimal ML-based lead position (patient #7).

The left panel demonstrates the ML-score map in a clinical responder (patient #2) with a 12% LVEF improvement. In this patient, almost the entire ML-score map is red, predicting a positive CRT response with any available LV site located at the lateral wall. Here, the referent and optimal pacing sites are located in adjacent LV segments and the maximum ML-score of 0.95 at ML-PS is slightly above the referent value of 0.94. So, this patient is predicted as a positive for CRT response (ML-score>0.5) at both the ref-PS and ML-PS. The LR prediction is in line with a considerable of LVEF improvement in this patient.

The central panel shows the ML-score map of a clinical non-responder (patient #1) with a 7% LVEF improvement. Here, the ML-scores at both the referent and optimal LV pacing sites are blue colored (0.14 and 0.27 < 0.5, respectively). Moreover, the overall map of ML-scores on the entire available LV surface is blue colored, predicting a low possible response to CRT in this patient. Correspondingly, this patient has a large postinfarction scar spreading over half of the LV segments (see the gray segments in LV AHA model).

The right panel shows the ML-score map of a clinical non-responder (patient #7) with a 6% LVEF. The patient was classified by the LR predictive model as a true negative at the ref-PS (ML-score=0.38, see the ref-PS located in the blue colour area on the ML-score map). At the same time, our algorithm predicts a narrow red area at the basal inferior LV segments where the patient is predicted as positive for CRT response, particularly at the optimal ML-PS position with maximum ML-score=0.77 (see ML-PS located in the red colour area on the ML-score map). Thus, our simulations suggest that this patient could possibly improve with the ML-based optimal pacing lead placement.

#### 3.3.1 Effect of pacing site position on ML-score

The first question we addressed in our study was the extent to which a change in LV pacing position has an impact on ML-score in a particular patient and in the overall population. Figure 4 shows ML-scores computed at multiple LV pacing sites tested for every patient. Here, patients were sorted according to their ML-score at ref-PS. In addition to the optimal ML-PS with maximum ML-score across the LV surface, two more LV pacing sites under BiV pacing were tested: LV pacing from the LAT area in baseline (LAT-PS) and an LV pacing site minimizing the total biventricular activation time (TAT-PS) as a measure of ventricular activation dyssynchrony. Apparently, variation in the ML-score was higher between the patients than within them. At the same time, the coefficient of variation of the ML-score within patient ranged from 0.1 to 1.31, indicating that in a number of patients the ML-score varied significantly with the position of the LV pacing site, which emphasizes the importance of pacing optimization.

**Figure 4.**
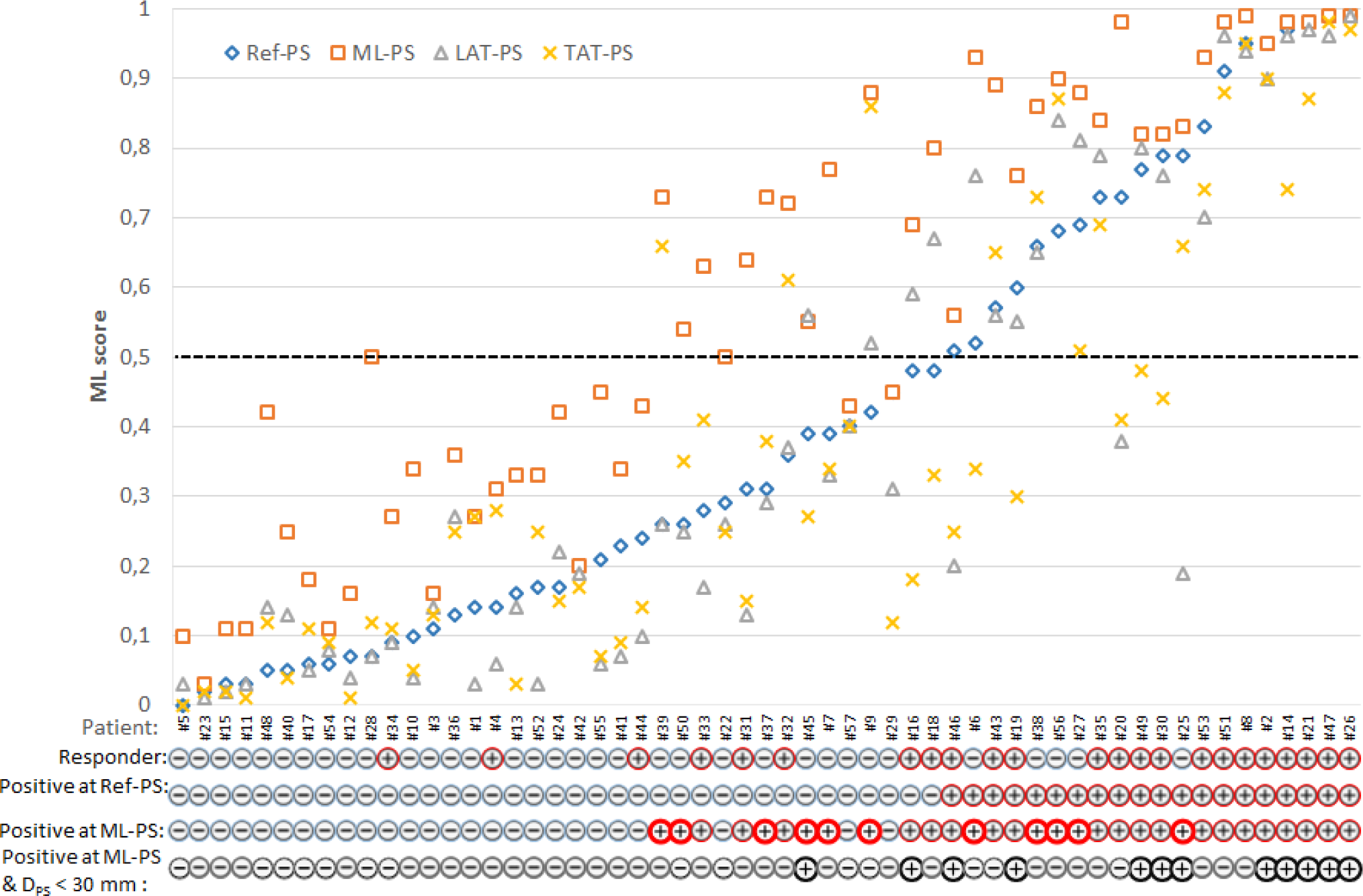
Individual ML-score for each patient with multiple LV pacing sites: clinical (reference) Ref-PS; optimal ML-PS, pacing at LV LAT area defined in baseline; pacing at the LV pacing site minimizing the total biventricular activation time - TAT-PS. Patients are sorted according to their ML-score at Ref-PS pacing. Following individual patient characteristics are shown below: a clinical responder (+) or non-responder (-), a positive (+)/negative (-) prediction of CRT response based on the ML-score value generated by the LR predictive model from the clinical data and simulations at Ref-PS and ML-PS, and a presence (+) of the combination of a positive ML-score at ML-PS and a low distance D_*PS*_<30 mm from the Ref-PS to optimal ML. Bold red circles indicate patients with a positive prediction of CRT response at ML-PS for the clinical non-responders.

The next question was whether the optimal ML-PS significantly raised the ML-score over the ref-PS. ML-score at ref-PS ranged from 0.03 to 0.99 between patients with a median of 0.31 [0.14,0.68] (see also Fig. 5). The ML-score at the optimal ML-PS ranged in the same interval, while the median was significantly higher 0,56 [0.33,0.87] (p<0.001, Fig. 5). The improvement in the optimal ML-score over the referent value ranging from 0 to 0.47 with a median of 0.16 [0.05, 0.25], and a high coefficient of variation of 1.64, demonstrate a substantial increase in ML-score in the majority of the patients. In particular, the optimal ML-score exceeded the reference value by 17[5;24]% in 89% of our patients (51 out of 57, 19 out of 23 responders and 32 out of 34 non-responders). The average optimal ML-score is significantly higher as compared to ref-PS in both the responder and non-responder groups (p<0.01, Table 1). At the same time, our model predicts a much higher relative increase (almost double) in ML-score at ML-PS in the non-responder group (see Table 1) with an increase of 18[8;26]% in 32 out of 34 non-responders.

**Figure 5.**
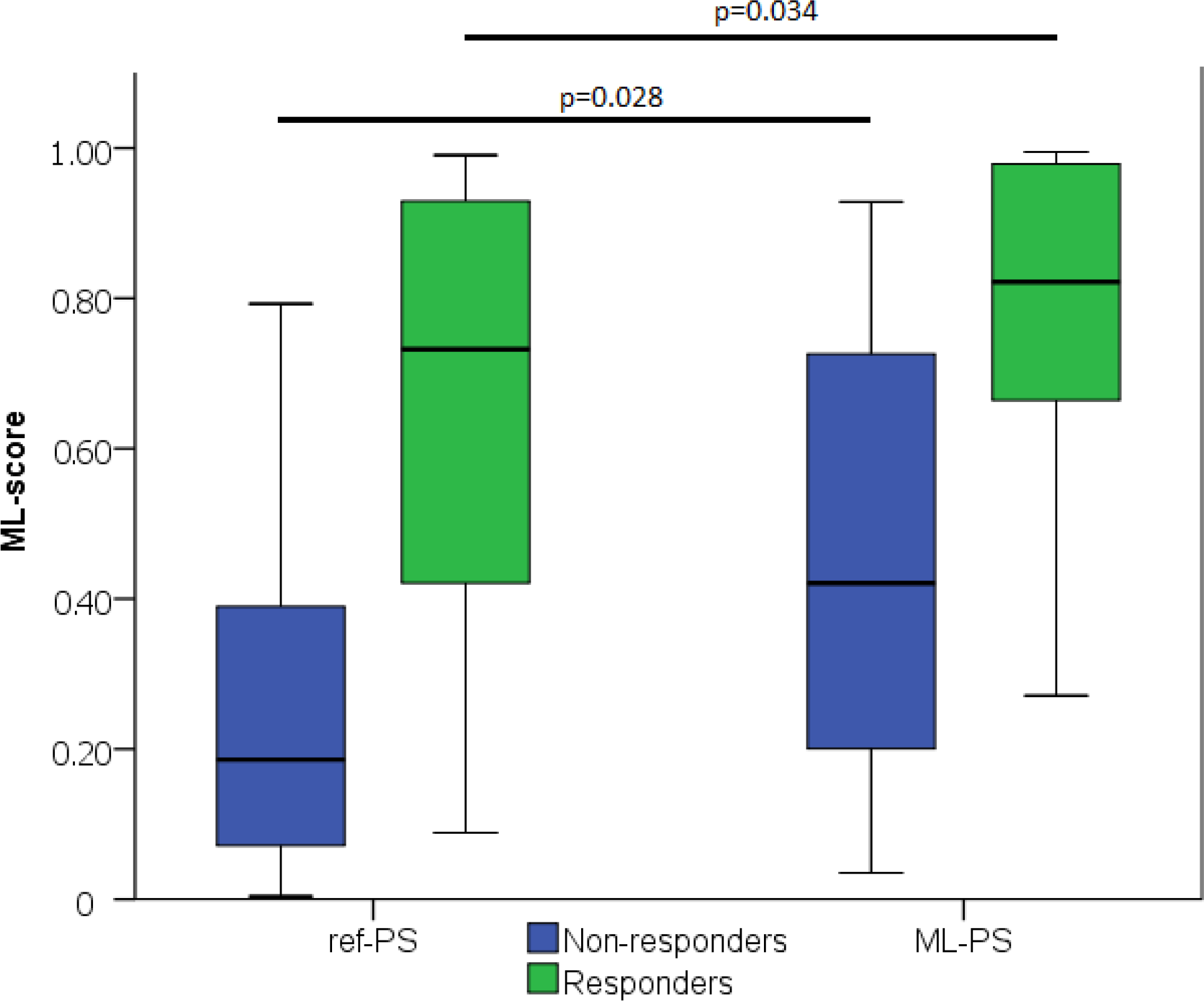
ML-score at ref-PS and ML-PS. Comparison between Responders and Non-responders (p) - two independent groups was carried out using Mann-Whitney test.

As defined above, the cut-off ML-score of 0.51 separates potential responders from non-responders. Comparing the ML-score at ML-PS and ref-PS, we found patients who were classified as negative (potential non-responders) with ref-PS but reclassified as positive (potential responders) with ML-PS. Figure 6 shows such upward transitions from the group of ML-score<0.5 at ref-PS to the group of ML-score>0.5 at ML-PS. There were eleven such transitions, shown in Table S7 in more detail. Here, five responders classified by the LR classifier as false negative at ref-PS move upward into the positive group at ML-PS (see +5 in the top left cell coming up from the bottom left cell). Moreover, 6 of 29 (21%) non-responders truly classified as negative at ref-PS are classified as positive at ML-PS (see +6 in the top right cell coming up from the bottom right cell). In total, according to the LR classifier the ratio of positive to negative for CRT response with an optimized ML-based pacing site increased considerably to 31-to-26 (54-to-46%) versus the ratio of 23-to-34 (40-to-60%) between responders and non-responders at ref-PS. At the same time, a few true clinical responders (3 out of 24) were still classified as false negative for CRT response even at the optimal LV pacing site ML-PS (Fig. 6, see 3 red dots in the right column with max ML-score < o.51 at ML-PS). Although it is difficult to identify specific factors contributing to the negative ML score value in the multifactorial LR model, we will discuss possible reasons for the false negative predictions in these patients below in the Discussion section #4.4).

**Figure 6.**
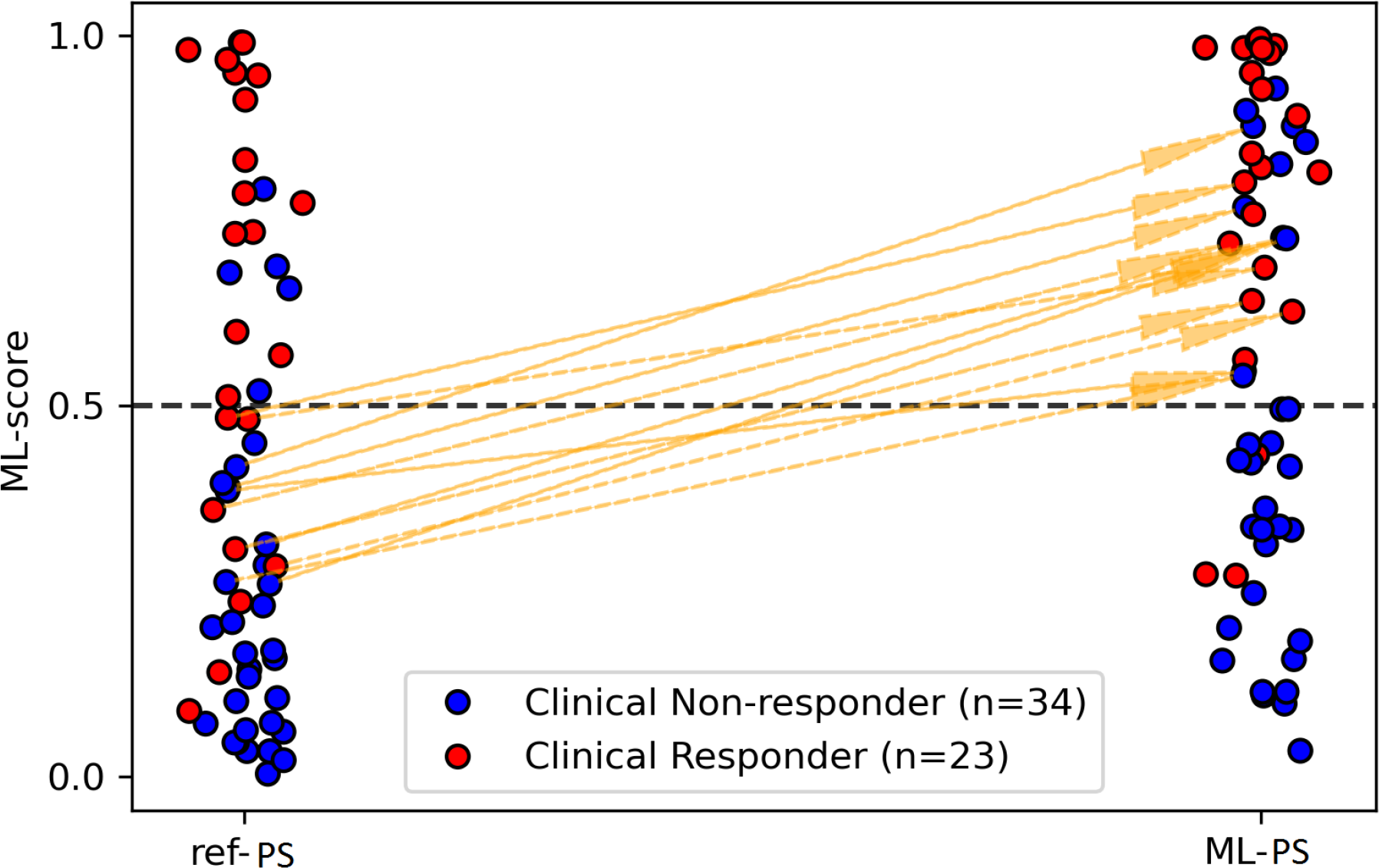
Transitions from negative to positive prediction of CRT response and reverse transitions when switching from ref-PS to ML-PS.

#### 3.3.2 Effects of optimal LV pacing site position on model-derived characteristics of ventricular activation

To explain the difference in the distributions of ML-scores depending on LV pacing site position, we compared the model-derived features characterising ventricular activation at ML-PS and ref-PS.

In a majority of models (52 (91%) out of the 57 models), BiV pacing from ML-PS was associated with a considerable decrease in all simulated indices characterizing ventricular activation dyssynchrony as compared with the baseline LBBB activation. The median TAT (99[88;111] ms) and QRSd (142[132;155] ms) at ML-PS are seen to be significantly shorter than at baseline (151[137;166] and 190[175;205], p<0.01, respectively). At the same time, no significant difference in average TAT and QRSd was found between ML-PS and ref-PS (Table 1).

Both the inter- and intra-ventricular dyssynchrony indices (RV-LV activation delay AD*_RV LV_* and ST-LV activation delay AD*_ST LV_*) in LBBB baseline have positive average values reflecting a significantly later activation of LV *versus* RV, and LV lateral wall *versus* septum (Table S5). Both indices reduce several times at ML-PS as compared with LBBB. However, no difference between ML-PS and ref-PS was found in the inter-ventricular dyssynchrony index AD*_RV LV_* . In contrast, the intra-ventricular dyssynchrony index AD*_ST LV_*, is slightly higher for ML-PS versus ref-PS. The positive AD*_ST LV_* (0.08[0.02;0.18]) at ML-PS suggests later activation of the LV lateral wall as compared to the septum, while the negative index (-0.09[-0.15;-0.03]) at ref-PS reflects later activation of the septum.

Similarly, there were no effects of pacing site position on TAT and QRSd, and on the dyssynchrony indices in the responder and non-responder groups. Thus, the peculiarities of any single model-derived indices tested under different pacing site positions could not explain the differences in the ML-score we found.

As described above, the Scar-LVPS distance from the LV pacing site to postinfarction scar area was selected as one of the significant model-driven features affecting LR classifier accuracy and the ML-score value. The Scar-LVPS distance was significantly longer at optimal ML-PS as compared to ref-PS (58[45;71] *vs* 34[10;49], respectively, p<0.05, Table 1). Moreover, a positive correlation (r=0.673, p<0.01) was found between the improvement in the ML-score and the extension of the Scar-LVPS distance when switching from ref-PS to ML-PS. Comparing the responder and non-responder groups, we observed that the Scar- LVPS distance was shorter for the non-responder group at ref-PS, while at ML-PS the distance increased significantly in the non-responder group, blurring the difference between the groups (Table 1). Finally, no correlation between the maximum ML-score and the Scar-LVPS distance was found at ML-PS.

### 3.4 Validation of LV pacing site optimization based on ML-score

Our current study utilized retrospective clinical data, with no patients treated according to the optimization procedure for LV lead implantation that we have developed. However, we were able to perform a virtual “clinical trial” of our optimization approach. First, we selected a group of patients classified as positive for CRT response with a maximum ML-score >0.5 (referred to as the “positive” group, n=31, 54% of the total 57 patients) identified by our optimised selection algorithm. Even with the use of an empirical approach to LV pacing lead implantation in patients from the selected positive group, the group contained 20 (65%) clinical responders and 11 (35%) non-responders paced from ref-PS. Thus, the clinical CRT response rate (65%) in the positive group selected according to our optimised approach was significantly higher than that of 40% in the entire cohort selected according to current guidelines.

In contrast, in the group classified as negative for CRT response (maximum ML-score<0.5, n=26, 46% of the total), there were only 3 (12%) clinical responders and 23 (88%) non-responders. Therefore, the odds ratio of becoming a responder with a positive prognosis versus a negative prognosis was 13.9 CI(3.4;57) in our cohort, suggesting a great potential of our approach to assist in the selection of CRT candidates.

Not surprisingly, a higher ΔLVEF was found in the positive group (14[8;17] *vs* 3[0;9]%, p<0.001) reflecting the higher ratio of responders against the negative group. However, no difference in ΔLVEF was found between positive and negative responders, nor between positive and negative non-responders. This fact suggests that if LV pacing site optimisation is not applied to selected patients, no additional LVEF improvement compared to the guidelines could be expected in positive candidates.

Next, to demonstrate the strength of LV pacing site optimisation, we classified our patients according to the proximity of the clinical ref-PS to the optimal ML-PS and assessed the significance of the distance (D_*PS*_) from the ref-PS to ML-PS as a measure of predicting CRT response. An average distance D_*PS*_ in the responders from the entire cohort was shorter against the non-responders (42*±*25 mm (median 35 [21,61] mm) versus 65*±*30 mm (median 64 [41,92] mm, p=0.005 respectively). The distinction in D*_PS_* between responders and non-responders was even higher in the positive group: 40*±*25 mm (median 29[20,61] mm) versus 66*±*28 mm (median 72[45,88] mm), p=0.005. Moreover, we found a moderate correlation (r=0,423, p<0,001) between the distance D_*PS*_ and ΔLVEF (Fig. S4, S5). The results suggested D_*PS*_ to be predictive of the CRT response.

Furthermore, we selected the two features - the maximum ML-score>0.5, and the distance D_*PS*_ - as measures to perform a linear discriminant analysis. The model yielded a ROC AUC of 0.85, p<0.001. The optimal cut-off point analysis showed that max ML-score=0.51 and D_*PS*_ = 30 mm divided the patients into responders and non-responders providing the best balance between sensitivity and specificity (sensitivity 87%, specificity 71%, positive predictive value 67%, and negative predictive value 89%).

In the group of patients with a maximum ML-score>0.5 (positive) and D_*PS*_<30 mm (n=12 (21%) out of total 57), the response rate was 83% (10 responders) which is much higher than 29% (13 responders out of 45) in the rest of the cohort. Selection of CRT candidates based on the positive prediction with maximum ML-score >0.5 and D_*PS*_<30 mm has the odds ratio for CRT response of 12.3 CI(2.4;64). Moreover, an average ΔLVEF value of 16*±*8 (median 15[11;20]) in this group is much higher than 7*±*8 (median 7[1;14])% (p<0.01) in the rest of the population, demonstrating a great improvement in the selected patients with a maximum ML-score >0.5 and LV pacing leads deployed in the proximity to the optimal LV pacing site.

The above results evidence a high potential of our ML-score based optimal LV lead placement to assist in selecting CRT candidates and guiding lead implantation.

## 4 DISCUSSION

### 4.1 ML-score based optimal LV lead position

In our recent study (26), we developed and validated ML classifiers to predict a long-term LVEF improvement of more than 10% in CRT recepients. In the current study, we used such an LR classifier as an essential component of a novel technology for optimising CRT. This technique may help to assess the probability of patient improvement before implantation, and to guide the procedure in selected patients. The CRT optimisation algorithm utilises characteristics of ventricular activation dyssynchrony derived from personalised computational models depending on the pacing site position. Here, we fixed the RV pacing site at the post-operative position in the BiV model settings because it did not vary between patients and was conventionally located in the RV apical region. The aim of this study was to evaluate the role of LV pacing site position and the ability to optimise CRT response by choice of position. The main advantage of using model simulations is the possibility of testing any accessible pacing site on the LV surface and predicting an optimal lead placement, which maximises the probability of patient improvement at BiV pacing.

As emphasised above, our approach to CRT optimisation can be applied during preoperative patient evaluation before data on RV-LV pacing sites and ECG recorded during BiV pacing are available for a given patient. The optimisation algorithm maximises an ML score of CRT response across the LV surface available for pacing using simulations of BiV pacing at different LV pacing site positions. Of note, a supervised ML classifier used to compute ML scores must be developed independently of and prior to the application of the optimisation procedure. At this stage, for the development of supervised LR classifiers, we need a training dataset of post-operative clinical data with determination of LV and RV pacing sites from CT images and determination of myocardial conductivity parameters based on clinically recorded QRSd at pacing. Since such a supervised LR classifier has been created (as a formulation with defined coefficients), one can use this classifier (with predefined and fixed coefficients) to compute ML scores for an arbitrary RV-LV lead setting using simulations of BiV pacing with that particular pacing site location. Of course, such an ML-classifier could be re-trained from time to time with an expanded dataset as new post-operative clinical data becomes available. However, since the ML classifier is set, no clinical pacing site location is required for our optimization algorithms.

The patient assessment algorithm involves building a personalised ventricular model; performing simulations with LBBB activation pattern and under BiV pacing with multiple LV pacing site positions; calculating ML-scores from the LR classifier fed with selected pre-implant clinical data and model-derived features, building a map of ML-score values over the entire LV surface, and finding the optimal LV pacing site position with a maximum ML-score using Gaussian process regression and Bayesian optimisation (Fig. 2). Ultimately, this approach gives a CRT operator an LV surface map with labelled areas for LV pacing, predicting either a positive (ML-score>0.5) or negative (ML-score<0.5) CRT response. Moreover, it identifies a target position for optimal LV pacing site that maximises the ML-score, predicting the highest possible probability of the patient’s response to CRT (see examples of ML-score maps in patient specific models in Figure 3). If the optimal ML-score is higher than cut-off value dividing responders and non-responders according to the ML classifier (maximum ML-score > 0.5), the patient may be considered as a candidate for CRT. The ML-based optimal LV lead position can be used to guide the implantation procedure.

We demonstrated a broad in-patient variation in the ML-score depending on the pacing site position (Figure 4). The ML-score varied more than 10-fold across the LV surface, and the range was much broader in the non-responder group of patients. One of the essential results of our study is that the ML-based optimal LV pacing site (ML-PS) provides the highest ML-score for CRT response in our patient population exceeding significantly the ML-score values at the clinical (ref-PS) and alternative pacing sites we tested. Moreover, in the non-responder group, the optimal ML-score showed a two-fold improvement compared to the referent value.

The strength of ML-based optimisation is clearly visualised in Figure 6. It shows a great number of transitions among the patients classified as negative for CRT response (ML-score < 0.5) at ref-PS into the positive group (ML-score > 0.5) at ML-PS. The maximum ML-score predicted a higher ratio of positive-to-negative patients (31/26 *≈* 1.2) as compared with the ratio of responders/nonresponders (23/34 *≈* 0.67).

### 4.2 On the regional distribution of optimal LV lead positions

Several clinical trials have recommended avoiding apical and anterior regions for LV pacing, where possible (3). Figure S1 compares the distribution of segments with LV pacing sites derived from the CT images with implanted leads and suggested by different optimisation approach. It can be seen that for ref-PS the lateral segments with LV pacing sites are more frequent (50 out of the 57 cases) in our population. For ML-based optimisation, lateral segments are more frequent (36 cases) as well, but both anterior (11 cases) and inferior (10 cases) segments are also representative in terms of the maximum ML-score. Analyzing the 11 cases with chosen optimal anterior segments for ML-PS, we found 4 responders with ref-PS in proximity to the optimal ML-PS (in the same or neighboring segments). The rest 7 non-responders were still predicted as negative at ML-PS (max ML-score < 0.5) suggesting a low likelihood of improvement. Thus, our model predictions are in line with clinical observations showing a small fraction of anterior segments among positive responses.

### 4.3 On the role of scarring area for optimal LV lead positioning

In our study, the extent of LV myocardial damage (both absolute and relative to the surviving myocardium volume) was not selected as a strong predictor of CRT response for the LR classifier. At the same time, the Scar-LVPS distance was selected as the third most important feature for CRT response prediction (see Fig. S3). It was selected by every feature selection algorithm we tested. This Scar-LVPS distance was the only model-driven feature that distinguished responders from non-responders in our population at the referent LV lead position (see Table 1), although, no correlation was found between the Scar-LVPS distance and LVEF improvement (r=0.18, p=0.211).

At the same time, we found a low positive correlation between the ML-score value and Scar-LVPS distance at ref-PS (r=0.419, p<0.001). In addition, we revealed a strong positive correlation between the improvement in the ML-score at the optimal ML-PS and the increase in Scar-LVPS distance against the ref-PS (r=0.673, p<0.01). The higher Scar-LVPS distances were associated with the maximum ML-scores (Table 1).

Our findings are consistent with the results of clinical studies which assessed the significance of postinfarction scar for CRT response. A higher LV dyssynchrony was shown to be strongly associated with echocardiographic response to CRT, while the total extent of scar derived from MRI and a match between the LV pacing site and transmural scar were found to be favourable to non-response (52). The location of scar in the posterolateral region of the LV, which is empirically thought to be a target site for LV lead implantation, was associated with lower response rates following CRT (8). Pezel and co-authors (7) found no difference in the presence and extent of scar between CRT responders and non-responders. However, in non-responders, the LV lead was more often over akinetic/dyskinetic regions. By contrast, the extent of the scar core and gray zone was automatically quantified using cardiac MRI analysis and the highest percentage of CRT response was observed in patients with low focal scar values and high QRS area before operation (6). However, the lack of direct MRI information regarding scarring in the cardiac tissue was mentioned as one of the limitations of the data they used. In our models we accounted for scar/fibrosis data and showed that this is essential for model predictions.

### 4.4 Validation of the optimal ML-PS approach

We analysed the benefits of our novel pacing site optimisation technology with respect to two clinical tasks: optimisation of patient selection and optimisation of procedure planning. The technology allows both steps to be performed preoperatively and simultaneously. Even without pacing site optimisation, a positive group of patients classified as favourable for CRT response by a maximum ML-score >0.5 contained a higher proportion of clinically proven responders with LVEF improvement greater than 10% (65% *vs* 40%) and showed a higher ΔLVEF (14[8;17] *vs* 9[2;15]%, p<0.01) compared to the entire cohort selected according to current guidelines. As expected, the ML-based positive group showed great advantages over the negative group with maximum ML-scores <0.5, consisting of only 12% of responders and showing a low ΔLVEF of 3[0;9] (p<0.01). These results demonstrate the high potential of our technology for patient selection for implantation.

Any predictive model has less than 100% sensitivity and specificity. Same with our ML based LR model, which is multifactorial, and a specific combination of input features contributes to the overall ML score output. Our optimized patient selection algorithm based on the positive response prediction from max ML-score >0.5 predicts clinical responders with a high sensitivity of 87% with 20 true positive (max Ml-score >0.5) and 3 false negative (max ML score <0.5) out of 23 true clinical responders. The 3 false negative patients #4,#34,#44 were ischemic cardiomyopathy patients with a high relative scar size in the myocardial tissue volume (31, 20 and 28% correspondingly) over the 75% quartile of the distribution 0.12 [0.07;0.23] and a great majority of damaged segments according to the AHA LV model (8, 10 and 7 damaged segments out of max 12 segments available for pacing). Two of the three patients have BMI of 31, 32 over the average 30 in non-responders. One of the latter has a high baseline LVEF of 32 over the mean 29 in non-responders. For two of the three patients, a short Scar-LVPS distance from the clinical LV pacing site to the scar area (0 and 16 mm) was lower than the 25% quartile for the distribution 44[19;53] in the responders. The above factors favor the negative ML-score prediction of the response for the patients, and it is actually difficult to distinguish particular features that are responsible for the false prediction in the multifactorial LR classifier. Despite max ML-score in these patients was 2-3 times higher than ML-score at ref-PS, it was still lower than the cutoff of 0.5. Note that in patient #43 the max ML-score was of 0.43, close to the responder threshold. We think that such patients with great extent of myocardial damage need more accurate scar area segmentation to get more specific response prognosis (see below Limitation Sec.).

Then, we demonstrated the validity of our optimisation approach as a procedure planning strategy. We showed that the distance D_*PS*_ from the clinical LV pacing position to the optimal site identified by our algorithm was an independent predictor of CRT response (ROC AUC = 0.72, p=0.005). The distance was shorter in the responders and correlated with LVEF improvement (Figure S4). Finally, discriminant analysis showed that a combination of the maximum ML-score and D_*PS*_ values is predictive of CRT response with a high accuracy (ROC AUC = 0.85, p<0.001). Together with max ML-score >0.5, a D_*PS*_ of less than 30 mm is suggested as a cut-off for predicting more than 10% LVEF improvement. The group of patients with maximum ML-scores >0.5 and D_*PS*_ <30 mm demonstrated an higher response rate (83% *vs* 29%, OR = 12.3 CL(2.4;64)) and a better LVEF improvement (15[11;20]% *vs* 7[1;14]%, p<0.01) as compared with the rest of the cohort.

The above results provide evidence of the high potential of our ML-score based LV pacing site optimisation for the selection of CRT candidates and for guiding lead implantation.

### 4.5 Which optimised LV lead position is better?

To compare the results of our ML-based optimisation approach, we tested two alternative LV pacing sites, which had been reported as potential for CRT improvement. In the first approach, we characterised the area of the LV late activation time (LAT) from the simulated ventricular activation map at baseline LBBB in each personalised model. The LAT pacing site (LAT-PS) was tested as an alternative LV pacing site during BiV pacing in each model. Using LAT-PS, we also calculated the distance D*_LAT −LV P S_* from the ML-PS and ref-LV to LAT area. In addition, we estimated the interventricular RV-LV delay as a time interval between the activation of the LV and RV pacing sites in the LBBB pattern. This feature is often used instead of the Q-LV delay measured from the onset of QRS complex to the activation time of the LV electrode in baseline or during RV pacing. Several studies have showed favourable effects of using the RV-LV delay or D*_LAT −LV P S_* as a guide (14, 25, 10, 53). By contrast, selecting the LAT pacing site in patients with non-LBBB was reported to have no benefits (54).

The other LV pacing alternative we tested involved LV pacing sites ensuring a minimum total biventricular activation time (TAT-PS). TAT is often used as a measure of ventricular dyssynchrony and its reduction via BiV pacing or at other pacing settings is considered as a target for stimulation design (24, 49, 55, 10). In clinical practice, direct assessment of both LAT area and TAT is complicated and requires invasive electrophysiological mapping to be performed. Currently, noninvasive body surface ECG mapping showed advantages in deriving ventricular activation characteristics (56, 55, 57). Personalised cardiac models present another useful tool for noninvasive prediction of the LAT area in LBBB and TAT values at various pacing configurations prior to the procedure (22).

In our patient cohort, we found no difference in the average RV-LV delay at ref-PS between the responders and non-responders (see Table S6). Neither was there a correlation between the RV-LV delay and LVEF improvement (r=-0.14, p=0.314). Similarly, there was no difference in the distance D*_LAT −LV P S_* between the responders and non-responders, and no correlation with LVEF improvement was found. Accordingly, in the univariate analysis these variables were not selected as independent predictors for CRT response; neither were they selected in the multivariate analysis. Therefore, none of these two features was used as an independent variable in our multivariable LR classifier of CRT response and in the algorithms of pacing site optimization.

As we showed above, TAT95 simulated in baseline LBBB was selected as one of the most important features for the multivariable LR classifier of CRT response, reflecting the significance of a wide QRSd in guiding patient selection for CRT. However, in consistency with clinical and simulated data on QRSd, no correlation was found between LVEF improvement and simulated TAT at either LBBB or BiV pacing. No correlation was found between LVEF improvement and relative reduction in TAT at BiV pacing against baseline for any pacing site strategy we tested (Table 1). The distance between TAT-PS and ref-PS did not differ between responders and non-responders and did not correlate with LVEF improvement either. Our simulation results are consistent with the simulation results by Lee et al. (25). These authors also found no significance of changes in several characteristics of ventricular activation dyssynchrtony in comparison to RV pacing as predictors of an acute hemodynamic response to BiV pacing.

The results of BiV pacing based on neither LAT-PS nor TAT-PS showed a more favourable prognosis of response to CRT compared to clinical ref-PS or optimal ML-PS. Despite the highest RV-LV delay at LAT-PS and the shortest TAT at TAT-PS as compared with other pacing sites (p<0.01), no improvement in the ML-scores over ref-PS was attained (see Figure S2 for the ML-score distribution at alternative pacing sites). The results suggest that any uniparametric strategy for targeting LV lead placement cannot improve CRT response prognosis. At the same time, the optimal ML-scores at ML-PS based on the multivariable LR predictions demonstrated the highest ML-score among the other pacing sites, suggesting the highest probability of CRT response.

The above results may seem inconsistent with several studies addressing pacing site optimization. Among the reasons of inconsistency could be difference in criteria used to define CRT response and the choice of either acute or chronic response for evaluating the outcome. In our current study, we trained predictive models on long-term LVEF improvement values during chronic BiV pacing in one year postimplant period. In line with other studies, we previously showed that the one-year period is optimal for assessing the magnitude of outcomes and benefits of therapy compared to and earlier follow-up (58). Over one year, the impact of the RV-LV delay could be less important. This suggestion is supported by our recent clinical observations (59). We compared two groups of patients with quadripolar LV leads: one group was paced according to maximum RV-LV delay, while in the other group a maximum RV-LV delay could not be approached during BiV pacing. We showed a faster improvement in the first group during the first 3-6 months after implantation. However, no difference was observed in LVEF improvement or ESV reduction in 12 months postimplant between the groups.

Another source for inter-study discrepancies could be related to data availability on the presence and distribution of LV scar or fibrosis and the accuracy of LAT area and TAT assessment. To our knowledge, our current study is the first one to use scar MRI data to predict LAT area and ventricular activation characteristics in models predicting response to CRT. Utilising both the distance D*_LAT −LV P S_* from the LV pacing site to the LAT area and the distance D*_Scar−LV P S_* from the LV pacing site to the scarring area, our analysis suggested the latter as a stronger predictor of pacing site optimisation.

## 5 STRENGTHS AND LIMITATIONS OF THE STUDY

In recent simulation studies, personalized cardiac models were used to reveal model-derived features correlating with CRT response (60, 61, 62, 63, 64). In two recent papers, model simulations were demonstrated to be predictive of LV pacing site optimization (24, 25). In our current study, we have developed an ML-based technique, using both clinical and simulated features. This technique provides an LV surface map predicting areas of positive and negative response and indicating the best possible place for LV lead guidance with a highest probability of CRT response for a given patient. Such a pre-operative virtual assistant can help select candidates for implantation and plan the procedure for selected patients. The results of our and other simulation studies demonstrate the potential of virtual clinical trials as a tool for exploring new approaches to CRT improvement.

In most studies, pacing site optimization is implemented either intra-procedurally with a parallel assessment of the acute response to BiV pacing, or post-implant in CRT device programming, particularly for quadripolar or multipace LV electrodes. Recently, the largest study using noninvasive 3D electrical activation mapping (ECGi) demonstrated the potential of using the ECGi technique in the pre-implantation planning strategy for CRT by enabling a noninvasive identification of the LAT area coupled to the identification of a suitable coronary vein (53). This approach is in line with our pre-implant optimization strategy, providing an operator with a target area for implantation. Note also that the reported strategies of pacing lead optimization are predominantly based on a single pre- or intra-operative feature (e.g. either LAT, or QRSd, or TAT, or *dP/dt_max_*, etc.). In contrast, our ML-based LV pacing site optimization accounts for multiple significant features related to the CRT response. A major added benefit of our approach is the use of model-derived predictions of BiV pacing from any available LV pacing site to estimate the probability of CRT response and to select an optimal pacing site that provides the highest probability. In addition, an essential advantage of our approach is the assessment of CRT efficacy prior to implantation, which can improve the response rate through optimal candidate selection and, together with optimised pacing, improve the outcomes.

There are several limitations in our study.

### Small dataset

In this study we had a limited dataset sampled from 57 patients. However, to the best of our knowledge, this is the largest model population used in simulation studies. Nevertheless, more data should be collected to split the datasets for training and testing the predictive models, in order to draw more robust conclusions. Still, more data are needed to validate the approach and confirm its usefulness for patient stratification and optimal lead guidance.

### Fibrosis/scar segmentation and simulation

We have shown a high importance of the distance D*_Scar−LV P S_* from the LV pacing site to the myocardial damage area for CRT optimization. In this study, we have simulated LV scarring area based on the labeling of damaged LV segments performed by the expert that analyzed the MRI scans. A more accurate segmentation of the raw MRI data should be used to confirm model predictions. In addition, our functional model of an infarct injury region was also simplified as we simulated the scar region as an inexcitable obstacle. Such a simplified assumption has often been used in modelling studies addressing the effects of the scar region on excitation propagation (see a review in (65)). However, experimental data in experimental animals and humans using MRI and optical mapping techniques showed a much more complex local structure and behaviour in the infarct zone (65, 66, 67, 68, 69, 70). The scar area, and particularly its border zone, has been shown to contain excitable inclusions of viable cells and fibroblast/myofibroblasts that influence the electrical activity in this area and in the surrounding myocardial tissue (68, 69). These structural and functional changes can result in a significant reduction in conduction velocity (but not complete absence of activity), which could be more accurately accounted for in the personalised models (65, 71, 72, 73).

### Purkinje network and LBBB level

Next, in this study we assumed complete LBBB when simulating baseline activation in the models. To simulate LBBB activation patterns in our models, we used a right branch of a synthetic model of the Purkinje network (74, 75, 76) with the left branch completely excluded. Previously, such Purkinje network models have been shown to reproduce an anatomical structure and myocardial activation patterns in the human and rabbit heart in norm and LBBB (76, 77). At the same time, there are experimental data on the morphological varieties of the Purkinje fibre network in mammalian hearts (78). However, currently it has not been possible to derive personalised data on the Purkinje network using non-invasive methods applicable in clinical practice. Furthermore, the effects of inter-subject variability in the Purkinje network on the ventricular activation pattern have not been addressed in detail in the experimental or simulation studies. Given that Purkinje myocardial junctions cover almost the entire endocardial surface of the right ventricular wall, we suggested that variations in the configuration of the right bundle branch should not significantly affect ventricular activation patterns in LBBB. Another facet of LBBB simulation is that current ECG criteria for LBBB do not distinguish between proximal and distal LBB conduction block, which may also affect the activation pattern in LV and requires further experimental and clinical consideration. In contrast, during BiV pacing, the onset of ventricular activation is defined by the location of RV-LV pacing sites, which we were able to define with high accuracy from CT images. In this case, the Purkinje network was excluded from the ventricular models and did not affect the results of BiV simulations.

### Model parameter tailoring

Another limitation of our personalised models was the use of an average QRSd and a global conductivity parameter for viable myocardium to fit the models to personal clinical ECG data. Such an approach provides a fast and robust solution, but may not reflect intra-ventricular variability in local conductivity, especially in areas where myocardial fibrosis/scar affects regional propagation.

We used different values of a global conductivity parameter in the LBBB and BiV models. The conductivity values were fitted from the clinical data on QRSd recorded pre- and post-operatively and reflected the clinically observed reduction in QRSd on pacing. The global parameter of myocardial conductivity in the Eikonal equation that we fitted to the QRSd value is actually an effective parameter of the model (not a purely physical value, unlike the local conductivity parameters) and depends on many factors such as LV end-diastolic volume, the relative volume of excitable myocardial tissue to nonexcitable tissue, the fraction of fibrotic tissue with reduced conductivity, the area of initial pacing and the location and proximity of the LV pacing lead to the infarct area, etc. These factors varied in the personalized models, which may explain a substantial variability in the global conductivity parameter (ranging from 0.2 to 1.5 for the scaling factor of the reference conductivity parameter value) that we identified from the clinical QRSd. Global conductivity parameters tended to be higher in BiV settings than in LBBB settings. This observation can be seen as a penalty for using only one parameter to compensate for many factors that contribute to the difference in activation time with BiV pacing compared to the baseline LBBB pattern. In particular, the increase in the global conductivity parameter was greater in BiV models with greater QRS reduction and a higher proportion of viable myocardium. In this study, we were not able to account for the postoperative change in ventricular geometry and LV volume, which decreases in a majority of patients with chronic pacing (especially in those we classified as responders), because we used postoperative CT scans for the ventricular geometry models used for both activation settings. It has recently been shown in a simulation study by Rodero and co-authors (24) that cardiac remodeling and reduction in ventricular volume after CRT implantation can reduce the effects of ventricular pacing. Then, our simplified models did not account for the mechanical activity, which is able to affect myocardial excitation depending on the contraction pattern and may contribute to the difference in ventricular dyssynchrony characteristics revealed by QRSd reduction. In addition, the timing of activation depends on the stimulation area, which we set in the BiV models as a small area of less than 2mm around the pacing site, whereas in reality this area could be larger depending on the stimulation settings used in patients. Moreover, myocardial conductivity during chronic pacing may itself change due to myocardial remodeling processes initiated by the change in activation sequence. Thus, an increase in global conductivity in the model at BiV settings could compensate for all of the above and other implicit factors to reproduce a substantial reduction in QRSd during postoperative myocardial remodeling. The difference in the global conductivity parameters we defined for LBBB and BiV models can be considered as a reflection of a number of uncertainties in the real system that are integrated in the single global parameter value.

### Model consistency and further improvements

Nevertheless, even with all the aforementioned assumptions and simplifications, we constructed and fitted our personalised models, which demonstrated the ability to capture clinical characteristics of ventricular activation at baseline and during BiV pacing and to reproduce clinically seen synchronisation of ventricular activation during BiV pacing (see Secs. 2.1 and 3.1 and Supplementary Fig. S6, S7, S8, S9, S10 on the consistency of model simulations to the clinical data). These results, together with the high accuracy of the ML classifier of CRT response utilizing the model-derived features at ref-PS, support the use of personalised models to predict the effects of BiV pacing within the ML-based approach we have developed to help CRT optimisation.

To ensure that the results of our study were robust to the global conductivity parameter used in the BiV settings, we compared the predictions of the optimal LV pacing site using BiV simulations with either the global conductivity parameter (BiV*_cond_*) defined from the postoperative QRSd at BiV pacing or the conductivity (LBBB*_cond_*) defined to match the preoperative clinical QRSd at baseline. The latter global conductivity parameter was used to compute simulated characteristics at both LBBB setting and BiV pacing with different tested LV pacing site locations. Corresponding ML scores were then computed using the predefined ML classifier we developed and used to predict an optimal LV lead position across the LV surface. We found that in 55 out of 57 (96%) patient models, the optimal lead position at the LBBB*_cond_* parameter was predicted in the same or neighboring segments of the LV AHA model as predicted from model simulations with the BiV*_cond_* parameter. The median distance between optimal LV pacing site locations was 3.9 mm, which is approximately the size of the stimulation area with the point source. In 75% of the models the distance was less than 10 mm and in 49 (86%) of the 57 models the distance was less than 20mm. The larger distance was observed in a few models with a large infarct size, where the optimal LV pacing location was predicted at the same distance from the infarct zone, but at the opposite position of the infarct area. Notably, ML-scores predicted from BiV simulations with different global conductivity parameters were not significantly different and predicted the same positive or negative CRT response for a patient. The only exception was observed in a non-responder model (patient #50), whose optimal ML-scores at different conductivities for BiV settings were close to each other (0.48 versus 0.52) and to the ML-score threshold of 0.5, which did not allow one to guarantee the positive response at the optimal position.

These observations allow us to suggest that, despite the limitations of the model, it is clearly able to predict the optimal position for the LV pacing site based on the only preoperative clinical data used for simulation at the BiV settings.

However, further improvements of personalised models could be performed in future to assess effects of more specific simulation of the Purkinje network morphology and LBBB level; scar/fibrosis area location, shape and texture; and tailoring personalised electrophysiological parameters based on the morphology of the QRS complex or the entire time-dependent ECG signal. Several approaches to the latter problem, assuming spatial heterogeneity of conductivity within the myocardial tissue and taking into account partially excitable areas of damaged myocardium, have recently been developed by several groups, including our team (79, 80), and could be implemented to fit model predictions to personal clinical data.

### Coronary sinus data merge

In this study, we identified a sort of ideal optimal LV pacing site on the entire LV surface. Here, we did not consider realistic anatomical constraints for epicardial LV lead access, which are limited by the available coronary sinus veins traditionally used for LV lead implantation. We plan to circumvent this limitation and validate our ML-based optimisation technique using ventricular models merged with coronary sinus visualisations. This will allow us to test the LV pacing sites within the coronary sinus approach and verify if our ML-based approach is able to improve CRT candidate selection and procedure outcome with more realistically optimised BiV pacing.

### Multi-modal clinical data

We have not addressed a number of strategies for patient selection and pacing site optimization that are becoming available with further development in ECGI and other imaging techniques. Recent studies have paid much attention to utilyzing vector ECG characteristics for CRT response prediction (81, 82, 83). The morphology and specific features of both conventional 12-lead ECG and vector ECG could be used to more accurately identify electrophysiological parameters in the personalised models (84, 85, 86) and to further improve predictive models of CRT response. In the present study, CRT response prediction involved simulated characteristics of ventricular activation and ECG derived from electrophysiological models. However, the synchronization of ventricular contraction and subsequent improvement in the mechanical performance of the ventricles is the main goal of the therapy. Recent studies have shown the predictive power of the mechanical indices that could be measured from CT or echocardiography images and accounted for in the predictive models of optimal pacing designs (25). Moreover, electromechanical models of cardiac activity (such as reported by (64, 63, 61, 62) and being developed by our team) could help perform direct simulations of LVEF, dP/dtmax changes and other mechanical biomarkers of CRT response, which can further improve ML based optimization of the CRT procedure.

### Alternative strategies for ventricular pacing

In this proof-of-concept study, we used our ML-based approach to optimize BiV pacing. Currently, there are data emerging on different pacing modality enhancing effects in selected patients depending on the ischemic or non-ischemic origin of HF, LBBB or non-LBBB activation, the levels of conduction system block, etc. New techniques for His-Purkinje and/or conduction system pacing, selective LV, RV or BiV, or multipole pacing, epicardial versus endocardial pacing and their possible combinations together with AV and VV delay optimisation for optimal ventricular fusion create a challenge for making the best choice (87, 88, 89). Only computational modeling provides a tool to test every possible combination and to suggest ones that help optimise patient outcome. The technology we have developed enables solving such complex problems.

Our approach enables one to account for more data available from various imaging modalities in the analysis and training of multivariable ML classifiers. Thus, the dataset for analysis can be expanded, CRT response criteria can be improved, and our general ML-based simulation approach to finding an optimal pacing configuration that maximises the probability of response can be further improved to accommodate data expansion. Furthermore, there is a general conclusion that can be drawn from the studies suggesting a strategy for LV pacing site optimisation to improve CRT response, that is, any such strategy needs to be evaluated prospectively in an intention-to-treat trial, in which data from different modalities can be included and different strategies can be compared.

## 6 CONCLUSION

We have developed algorithms to identify an optimal LV pacing position across the accessible LV surface that maximizes the ML-score generated by the LR classifier of CRT response.

In the group of selected patients with maximum ML values > 0.5 predictive of a positive CRT response, the response rate improved compared to the negative group and the overall cohort.

The proximity of implanted LV leads to the optimal pacing site, which can be determined non-invasively and preoperatively using our optimisation technique, was shown to be a strong independent predictor of CRT response.

Our non-invasive approach to identifying a maximum ML score of CRT response can help improve patient selection for CRT. At the same time, the best pacing site location predicted by our algorithms can be used to guide lead placement. Overall, our approach may help improve the efficacy of CRT.

## AUTHORS CONTRIBUTION

OS, AD, TC, AB, and SK participated in conceptualisation and design of the study, data collection, processing, and analysis. SZ, TL, VL and DL performed patient selection and treatment, clinical data collection and preprocessing. AB and SZ performed segmentation CT and MRI images and construction of anatomical models. AD, SK, AB performed personalised computational simulations of myocardial electrical activation. AD implemented algorithms of the pacing site optimisation. SK performed machine learning and built classifiers. TC contributed to the statistical analysis and data interpretation. OS supervised all stages of study execution and data analysis, and was a major contributor to writing the manuscript. All authors contributed to manuscript preparation and approved the final version of the manuscript.

## Data Availability

All data produced in the present study are available upon reasonable request to the authors

## REFERENCES

1 Daubert C, Behar N, Martins RP, Mabo P, Leclercq C. Avoiding non-responders to cardiac resynchronization therapy: A practical guide 38 (2017) 1463–1472. doi:10.1093/eurheartj/ehw270.

2 Wouters PC, Vernooy K, Cramer MJ, Prinzen FW, Meine M. Optimizing lead placement for pacing in dyssynchronous heart failure: The patient in the lead. Heart Rhythm 18 (2021) 1024–1032. doi:10.1016/j.hrthm.2021.02.011.

3 Butter C, Georgi C, Stockburger M. Optimal CRT Implantation—Where and How To Place the Left-Ventricular Lead? 18 (2021) 329–344. doi:10.1007/s11897-021-00528-9.

4 Mullens W, Auricchio A, Martens P, Witte K, Cowie MR, Delgado V, et al. Optimized implementation of cardiac resynchronization therapy: a call for action for referral and optimization of care: A joint position statement from the Heart Failure Association (HFA), European Heart Rhythm Association (EHRA), and European Association of Cardiovascular Imaging (EACVI) of the European Society of Cardiology. European Journal of Heart Failure 22 (2020) 2349–2369. doi:10.1002/ejhf.2046.

5 Sieniewicz BJ, Gould J, Porter B, Sidhu BS, Behar JM, Claridge S, et al. Optimal site selection and image fusion guidance technology to facilitate cardiac resynchronization therapy. Expert Review of Medical Devices 15 (2018) 555–570. doi:10.1080/17434440.2018.1502084.

6 Nguyên UC, Claridge S, Vernooy K, Engels EB, Razavi R, Rinaldi CA, et al. Relationship between vectorcardiographic QRSarea, myocardial scar quantification, and response to cardiac resynchronization therapy. Journal of Electrocardiology 51 (2018) 457–463. doi:10.1016/J.JELECTROCARD.2018.01.009.

7 Pezel T, Mika D, Logeart D, Cohen-Solal A, Beauvais F, Henry P, et al. Characterization of non-response to cardiac resynchronization therapy by post-procedural computed tomography. PACE - Pacing and Clinical Electrophysiology 44 (2021) 135–144. doi:10.1111/pace.14134.

8 Chalil S, Foley PW, Muyhaldeen SA, Patel KC, Yousef ZR, Smith RE, et al. Late gadolinium enhancement-cardiovascular magnetic resonance as a predictor of response to cardiac resynchronization therapy in patients with ischaemic cardiomyopathy. Europace 9 (2007) 1031–1037. doi:10.1093/europace/eum133.

9 Sommer A, Kronborg MB, Nørgaard BL, Stephansen C, Poulsen SH, Kristensen J, et al. Longer inter-lead electrical delay is associated with response to cardiac resynchronization therapy in patients with presumed optimal left ventricular lead position. Europace 20 (2018) 1630–1637. doi:10.1093/europace/eux384.

10 Fyenbo DB, Sommer A, Nørgaard BL, Kronborg MB, Kristensen J, Gerdes C, et al. Long-term outcomes in a randomized controlled trial of multimodality imaging-guided left ventricular lead placement in cardiac resynchronization therapy. EP Europace 24 (2022) 828–834. doi:10.1093/europace/euab314.

11 Varma N, O’Donnell D, Bassiouny M, Ritter P, Pappone C, Mangual J, et al. Programming cardiac resynchronization therapy for electrical synchrony: Reaching beyond left bundle branch block and left ventricular activation delay. Journal of the American Heart Association 7 (2018) 1–12. doi:10.1161/JAHA.117.007489.

12 Strik M, Ploux S, Huntjens PR, Nguyên UC, Frontera A, Eschalier R, et al. Response to cardiac resynchronization therapy is determined by intrinsic electrical substrate rather than by its modification. International Journal of Cardiology 270 (2018) 143–148. doi:10.1016/j.ijcard.2018.06.005.

13 Haqqani HM, Burri H, Kayser T, Carter N, Gold MR. Association of interventricular activation delay with clinical outcomes in cardiac resynchronization therapy. Heart Rhythm (2022). doi:10.1016/j.hrthm.2022.11.012.

14 Gold MR, Yu Y, Wold N, Day JD. The role of interventricular conduction delay to predict clinical response with cardiac resynchronization therapy. Heart Rhythm 14 (2017) 1748–1755. doi:10.1016/j.hrthm.2017.10.016.

15 Kalscheur MM, Kipp RT, Tattersall MC, Mei C, Buhr KA, Demets DL, et al. Machine Learning Algorithm Predicts Cardiac Resynchronization Therapy Outcomes: Lessons from the COMPANION Trial. Circulation: Arrhythmia and Electrophysiology 11 (2018). doi:10.1161/CIRCEP.117.005499.

16 Tokodi M, Schwertner WR, Kovács A, Tosér Z, Staub L, Sárkány A, et al. Machine learning-based mortality prediction of patients undergoing cardiac resynchronization therapy: the SEMMELWEIS-CRT score. European Heart Journal 41 (2020) 1747–1756. doi:10.1093/eurheartj/ehz902.

17 Tokodi M, Behon A, Merkel ED, Kovács A, Tosér Z, Sárkány A, et al. Sex-specific patterns of mortality predictors among patients undergoing cardiac resynchronization therapy: A machine learning approach. Frontiers in Cardiovascular Medicine 8 (2021) 87. doi:10.3389/fcvm.2021.611055.

18 Hu SY, Santus E, Forsyth AW, Malhotra D, Haimson J, Chatterjee NA, et al. Can machine learning improve patient selection for cardiac resynchronization therapy? PLOS ONE 14 (2019) 1–13. doi:10.1371/journal.pone.0222397.

19 Cikes M, Sanchez-Martinez S, Claggett B, Duchateau N, Piella G, Butakoff C, et al. Machine learning-based phenogrouping in heart failure to identify responders to cardiac resynchronization therapy. European Journal of Heart Failure 21 (2019) 74–85. doi:https://doi.org/10.1002/ejhf.1333.

20 Feeny AK, Rickard J, Trulock KM, Patel D, Toro S, Moennich LA, et al. Machine learning of 12- lead qrs waveforms to identify cardiac resynchronization therapy patients with differential outcomes. Circulation: Arrhythmia and Electrophysiology 13 (2020) e008210. doi:10.1161/CIRCEP.119.008210.

21 Feeny AK, Rickard J, Patel D, Toro S, Trulock KM, Park CJ, et al. Machine learning prediction of response to cardiac resynchronization therapy. Circulation: Arrhythmia and Electrophysiology 12 (2019) e007316. doi:10.1161/CIRCEP.119.007316.

22 Lee A, Nguyen U, Razeghi O, Gould J, Sidhu B, Sieniewicz B, et al. A rule-based method for predicting the electrical activation of the heart with cardiac resynchronization therapy from non-invasive clinical data. Medical Image Analysis 57 (2019) 197–213. doi:https://doi.org/10.1016/j.media.2019.06.017.

23 Villongco CT, Krummen DE, Omens JH, McCulloch AD. Non-invasive, model-based measures of ventricular electrical dyssynchrony for predicting CRT outcomes. EP Europace 18 (2016) iv104–iv112. doi:10.1093/europace/euw356.

24 Rodero C, Strocchi M, Lee AW, Rinaldi CA, Vigmond EJ, Plank G, et al. Impact of anatomical reverse remodelling in the design of optimal quadripolar pacing leads: A computational study. Computers in Biology and Medicine 140 (2022) 105073. doi:10.1016/j.compbiomed.2021.105073.

25 Lee AW, Razeghi O, Solis-Lemus JA, Strocchi M, Sidhu B, Gould J, et al. Non-invasive simulated electrical and measured mechanical indices predict response to cardiac resynchronization therapy. Computers in Biology and Medicine 138 (2021) 104872. doi:10.1016/j.compbiomed.2021.104872.

26 Khamzin S, Dokuchaev A, Bazhutina A, Chumarnaya T, Zubarev S, Lyubimtseva T, et al. Machine learning prediction of cardiac resynchronisation therapy response from combination of clinical and model-driven data. Frontiers in Physiology 12 (2021). doi:10.3389/fphys.2021.753282.

27 Arevalo HJ, Vadakkumpadan F, Guallar E, Jebb A, Malamas P, Wu KC, et al. Arrhythmia risk stratification of patients after myocardial infarction using personalized heart models. Nature communications 7 (2016) 11437.

28 Popescu DM, Shade JK, Lai C, Aronis KN, Ouyang D, Moorthy MV, et al. Arrhythmic sudden death survival prediction using deep learning analysis of scarring in the heart. Nature Cardiovascular Research 1 (2022) 334–343.

29 Shade JK, Prakosa A, Popescu DM, Yu R, Okada DR, Chrispin J, et al. Predicting risk of sudden cardiac death in patients with cardiac sarcoidosis using multimodality imaging and personalized heart modeling in a multivariable classifier. Science Advances 7 (2021) 8020–8048. doi:10.1126/SCIADV.ABI8020.

30 Shade JK, Ali RL, Basile D, Popescu D, Akhtar T, Marine JE, et al. Preprocedure application of machine learning and mechanistic simulations predicts likelihood of paroxysmal atrial fibrillation recurrence following pulmonary vein isolation. Circulation: Arrhythmia and Electrophysiology 13 (2020) 617–627. doi:10.1161/CIRCEP.119.008213.

31 Bayer JD, Blake RC, Plank G, Trayanova NA. A novel rule-based algorithm for assigning myocardial fiber orientation to computational heart models. Annals of Biomedical Engineering 40 (2012) 2243– 2254. doi:10.1007/s10439-012-0593-5.

32 Arevalo H, Plank G, Helm P, Halperin H, Trayanova N. Tachycardia in post-infarction hearts: Insights from 3d image-based ventricular models. PLOS ONE 8 (2013) 1–10. doi:10.1371/journal.pone.0068872.

33 Lopez-Perez A, Sebastian R, Izquierdo M, Ruiz R, Bishop M, Ferrero JM. Personalized cardiac computational models: from clinical data to simulation of infarct-related ventricular tachycardia. Frontiers in physiology 10 (2019) 580.

34 Mangileva D, Konovalov P, Dokuchaev A, Solovyova O, Panfilov AV. Period of arrhythmia anchored around an infarction scar in an anatomical model of the human ventricles. Mathematics 9 (2021) 2911.

35 Keener JP. An eikonal-curvature equation for action potential propagation in myocardium. Journal of Mathematical Biology 29 (1991) 629–651. doi:10.1007/BF00163916.

36 Franzone PC, Guerri L. Spreading of excitation in 3-d models of the anisotropic cardiac tissue. validation of the eikonal model. Mathematical Biosciences 113 (1993) 145–209. doi:10.1016/0025-5564(93)90001-Q.

37 Pezzuto S, Kal’avský P, Potse M, Prinzen FW, Auricchio A, Krause R. Evaluation of a rapid anisotropic model for ecg simulation. Frontiers in Physiology 0 (2017) 265. doi:10.3389/FPHYS.2017.00265.

38 Pullan AJ, Tomlinson KA, Hunter PJ. A finite element method for an eikonal equation model of myocardial excitation wavefront propagation. http://dx.doi.org/10.1137/S0036139901389513 **63** (2006) 324–350. doi:10.1137/S0036139901389513.

39 Camps J, Lawson B, Drovandi C, Minchole A, Wang ZJ, Grau V, et al. Inference of ventricular activation properties from non-invasive electrocardiography. Medical Image Analysis 73 (2021) 102143. doi:10.1016/j.media.2021.102143.

40 Pezzuto S, Prinzen FW, Potse M, Maffessanti F, Regoli F, Caputo ML, et al. Reconstruction of three-dimensional biventricular activation based on the 12-lead electrocardiogram via patient-specific modelling. Europace 23 (2021) 640–647. doi:10.1093/europace/euaa330.

41 ten Tusscher KH.J. Alternans and spiral breakup in a human ventricular tissue model. AJP: Heart and Circulatory Physiology 291 (2006) H1088–H1100. doi:10.1152/ajpheart.00109.2006.

42 Sahli Costabal F, Hurtado DE, Kuhl E. Generating Purkinje networks in the human heart. Journal of Biomechanics 49 (2016) 2455–2465. doi:10.1016/j.jbiomech.2015.12.025.

43 Plesinger F, van Stipdonk AM, Smisek R, Halamek J, Jurak P, Maass AH, et al. Fully automated QRS area measurement for predicting response to cardiac resynchronization therapy. Journal of Electrocardiology 63 (2020) 159–163. doi:10.1016/j.jelectrocard.2019.07.003.

44 Yin R, Liu Z, Zheng N. A simulation-based model for continuous network design problem using bayesian optimization. IEEE Transactions on Intelligent Transportation Systems 23 (2022) 20352– 20367.

45 Frazier PI. A tutorial on bayesian optimization. *arXiv preprint arXiv:1807.02811* (2018).

46 Williams CK, Rasmussen CE. Gaussian processes for machine learning, vol. 2 (MIT press Cambridge, MA) (2006).

47 Sommer A, Kronborg MB, Nørgaard BL, Poulsen SH, Bouchelouche K, Böttcher M, et al. Multimodality imaging-guided left ventricular lead placement in cardiac resynchronization therapy: a randomized controlled trial. European Journal of Heart Failure 18 (2016) 1365–1374. doi:10.1002/ejhf.530.

48 Yagishita D, Shoda M, Yagishita Y, Ejima K, Hagiwara N. Time interval from left ventricular stimulation to QRS onset is a novel predictor of nonresponse to cardiac resynchronization therapy. Heart Rhythm 16 (2019) 395–402. doi:10.1016/j.hrthm.2018.08.035.

49 Pereira H, Jackson TA, Claridge S, Behar JM, Yao C, Sieniewicz B, et al. Comparison of Echocardiographic and Electrocardiographic Mapping for Cardiac Resynchronisation Therapy Optimisation. Cardiology Research and Practice 2019 (2019). doi:10.1155/2019/4351693.

50 Logg A, Wells GN. Dolfin: Automated finite element computing. ACM Trans. Math. Softw. 37 (2010). doi:10.1145/1731022.1731030.

51 Bingham E, Chen JP, Jankowiak M, Obermeyer F, Pradhan N, Karaletsos T, et al. Pyro: Deep Universal Probabilistic Programming. Journal of Machine Learning Research (2018).

52 Marsan NA, Westenberg JJ, Ypenburg C, van Bommel RJ, Roes S, Delgado V, et al. Magnetic resonance imaging and response to cardiac resynchronization therapy: relative merits of left ventricular dyssynchrony and scar tissue. European Heart Journal 30 (2009) 2360–2367. doi:10.1093/EURHEARTJ/EHP280.

53 Parreira L, Tsyganov A, Artyukhina E, Vernooy K, Tondo C, Adragao P, et al. Non-invasive 3d electrical activation mapping to predict crt response: site of latest lv activation relative to pacing site. Europace (in press).

54 Singh JP, Berger RD, Doshi RN, Lloyd M, Moore D, Stone J, et al. Targeted Left Ventricular Lead Implantation Strategy for Non-Left Bundle Branch Block Patients: The ENHANCE CRT Study. JACC: Clinical Electrophysiology 6 (2020) 1171–1181. doi:10.1016/j.jacep.2020.04.034.

55 Zweerink A, Zubarev S, Bakelants E, Potyagaylo D, Stettler C, Chmelevsky M, et al. His-Optimized Cardiac Resynchronization Therapy With Ventricular Fusion Pacing for Electrical Resynchronization in Heart Failure. JACC: Clinical Electrophysiology 7 (2021) 881–892. doi:10.1016/j.jacep.2020.11.029.

56 Sieniewicz BJ, Jackson T, Claridge S, Pereira H, Gould J, Sidhu B, et al. Optimization of CRT programming using non-invasive electrocardiographic imaging to assess the acute electrical effects of multipoint pacing. Journal of Arrhythmia 35 (2019) 267–275. doi:10.1002/joa3.12153.

57 Sedova K, Repin K, Donin G, Van Dam P, Kautzner J. Clinical utility of body surface potential mapping in CRT patients. Arrhythmia and Electrophysiology Review 10 (2021) 113–119. doi:10.15420/aer.2021.14.

58 Chumarnaya TV, Lyubimtseva TA, Solodushkin SI, Lebedeva VK, Lebedev DS, Solovieva OE. Evaluation of the long-term effectiveness of cardiac resynchronization therapy. Russian Journal of Cardiology 26 (2021) 48–60. doi:10.15829/1560-4071-2021-4531.

59 Chumarnaya TV, Lyubimtseva TA, Lebedeva VK, Gasimova NZ, Lebedev DS, Solovieva OE. Evaluation of interventricular delay during cardiac resynchronization therapy in patients with quadripolar systems in long-term postoperative follow-up. Russian Journal of Cardiology 27 (2022) 60–69. doi:10.15829/1560-4071-2022-5121.

60 Villongco CT, Krummen DE, Omens JH, McCulloch AD. Non-invasive, model-based measures of ventricular electrical dyssynchrony for predicting CRT outcomes. Europace : European pacing, arrhythmias, and cardiac electrophysiology : journal of the working groups on cardiac pacing, arrhythmias, and cardiac cellular electrophysiology of the European Society of Cardiology 18 (2016) iv104–iv112. doi:10.1093/europace/euw356.

61 Lee AW., Mendonca Costa C, Strocchi M, Rinaldi CA, Niederer SA. Computational Modeling for Cardiac Resynchronization Therapy. Journal of Cardiovascular Translational Research 11 (2018) 92–108. doi:10.1007/s12265-017-9779-4.

62 Sermesant M, Chabiniok R, Chinchapatnam P, Mansi T, Billet F, Moireau P, et al. Patient-specific electromechanical models of the heart for the prediction of pacing acute effects in CRT: A preliminary clinical validation. Medical Image Analysis 16 (2012) 201–215. doi:10.1016/j.media.2011.07.003.

63 Okada Ji, Washio T, Nakagawa M, Watanabe M, Kadooka Y, Kariya T, et al. Multi-scale, tailor-made heart simulation can predict the effect of cardiac resynchronization therapy. Journal of Molecular and Cellular Cardiology 108 (2017) 17–23. doi:10.1016/j.yjmcc.2017.05.006.

64 Isotani A, Yoneda K, Iwamura T, Watanabe M, ichi Okada J, Washio T, et al. Patient-specific heart simulation can identify non-responders to cardiac resynchronization therapy. Heart and Vessels 35 (2020) 1135–1147. doi:10.1007/s00380-020-01577-1.

65 Connolly AJ, Bishop MJ. Computational representations of myocardial infarct scars and implications for arrhythmogenesis. Clinical Medicine Insights: Cardiology 10s1 (2016) CMC.S39708. doi:10.4137/CMC.S39708. PMID: 27486348.

66 Ashikaga H, Sasano T, Dong J, Zviman MM, Evers R, Hopenfeld B, et al. Magnetic resonance– based anatomical analysis of scar-related ventricular tachycardia: implications for catheter ablation. Circulation research 101 (2007) 939–947.

67 Richardson WJ, Clarke SA, Quinn TA, Holmes JW. Physiological implications of myocardial scar structure. Comprehensive Physiology 5 (2015) 1877.

68 Rutherford SL, Trew ML, Sands GB, LeGrice IJ, Smaill BH. High-resolution 3-dimensional reconstruction of the infarct border zone: impact of structural remodeling on electrical activation. Circulation research 111 (2012) 301–311.

69 Lodrini AM, Goumans MJ. Cardiomyocytes cellular phenotypes after myocardial infarction. Frontiers in cardiovascular medicine (2021) 1629.

70 Martínez MS, García A, Luzardo E, Chávez-Castillo M, Olivar LC, Salazar J, et al. Energetic metabolism in cardiomyocytes: molecular basis of heart ischemia and arrhythmogenesis. Vessel Plus 1 (2017) 130–141.

71 Dokuchaev A, Panfilov AV, Solovyova O. Myocardial fibrosis in a 3d model: effect of texture on wave propagation. Mathematics 8 (2020) 1352.

72 Mendonca Costa C, Plank G, Rinaldi CA, Niederer SA, Bishop MJ. Modeling the electrophysiological properties of the infarct border zone. Frontiers in physiology 9 (2018) 356.

73 Ringenberg J, Deo M, Filgueiras-Rama D, Pizarro G, Ibañez B, Peinado R, et al. Effects of fibrosis morphology on reentrant ventricular tachycardia inducibility and simulation fidelity in patient-derived models. Clinical Medicine Insights: Cardiology 8s1 (2014) CMC.S15712. doi:10.4137/CMC.S15712. PMID: 25368538.

74 Sebastian R, Zimmerman V, Romero D, Frangi AF. Construction of a computational anatomical model of the peripheral cardiac conduction system. IEEE Transactions on Biomedical Engineering 58 (2011) 3479–3482.

75 Ijiri T, Ashihara T, Yamaguchi T, Takayama K, Igarashi T, Shimada T, et al. A procedural method for modeling the purkinje fibers of the heart. The journal of physiological sciences 58 (2008) 481–486.

76 Barber F, Langfield P, Lozano M, García-Fernández I, Duchateau J, Hocini M, et al. Estimation of personalized minimal purkinje systems from human electro-anatomical maps. IEEE Transactions on Medical Imaging 40 (2021) 2182–2194.

77 Zhu H, Jin L, Huang Y, Wu X. A computer simulation research of two types of cardiac physiological pacing. Applied Sciences 11 (2021) 449.

78 Ono N, Yamaguchi T, Ishikawa H, Arakawa M, Takahashi N, Saikawa T, et al. Morphological varieties of the purkinje fiber network in mammalian hearts, as revealed by light and electron microscopy. Archives of histology and cytology 72 (2009) 139–149.

79 Albors C, Lluch Gomez JF, Cedilnik N, Mountris KA, Mansi T, et al. Meshless electrophysiological modeling of cardiac resynchronization therapymdash;benchmark analysis with finite-element methods in experimental data. Applied Sciences 12 (2022).

80 Moreau-Villéger V, Delingette H, Sermesant M, Ashikaga H, McVeigh E, Ayache N. Building maps of local apparent conductivity of the epicardium with a 2-d electrophysiological model of the heart. IEEE Transactions on Biomedical Engineering 53 (2006) 1457–1466.

81 Van Deursen CJ, Vernooy K, Dudink E, Bergfeldt L, Crijns HJ, Prinzen FW, et al. Vectorcardiographic QRS area as a novel predictor of response to cardiac resynchronization therapy. Journal of Electrocardiology 48 (2015) 45–52. doi:10.1016/j.jelectrocard.2014.10.003.

82 Emerek K, Friedman DJ, Sørensen PL, Hansen SM, Larsen JM, Risum N, et al. Vectorcardiographic QRS area is associated with long-term outcome after cardiac resynchronization therapy. Heart Rhythm 16 (2019) 213–219. doi:10.1016/j.hrthm.2018.08.028.

83 Ghossein MA, van Stipdonk AM, Prinzen FW, Vernooy K. Vectorcardiographic QRS area as a predictor of response to cardiac resynchronization therapy. Journal of Geriatric Cardiology 19 (2022) 9–20. doi:10.11909/j.issn.1671-5411.2022.01.003.

84 Gillette K, Gsell MA, Prassl AJ, Karabelas E, Reiter U, Reiter G, et al. A framework for the generation of digital twins of cardiac electrophysiology from clinical 12-leads ecgs. Medical Image Analysis 71 (2021) 102080.

85 Pezzuto S, Prinzen FW, Potse M, Maffessanti F, Regoli F, Caputo ML, et al. Reconstruction of three-dimensional biventricular activation based on the 12-lead electrocardiogram via patient-specific modelling. EP Europace 23 (2021) 640–647.

86 Costa CM, Gemmell P, Elliott MK, Whitaker J, Campos FO, Strocchi M, et al. Determining anatomical and electrophysiological detail requirements for computational ventricular models of porcine myocardial infarction. Computers in Biology and Medicine 141 (2022) 105061.

87 Mariani MV, Piro A, Forleo GB, Della Rocca DG, Natale A, Miraldi F, et al. Clinical, procedural and lead outcomes associated with different pacing techniques: a network meta-analysis. International Journal of Cardiology (2023).

88 Rijks J, Ghossein MA, Wouters PC, Dural M, Maass AH, Meine M, et al. Comparison of the relation of the esc 2021 and esc 2013 definitions of left bundle branch block with clinical and echocardiographic outcome in cardiac resynchronization therapy. Journal of Cardiovascular Electrophysiology (2023).

89 Zweerink A, Zubarev S, Bakelants E, Potyagaylo D, Stettler C, Chmelevsky M, et al. His-optimized cardiac resynchronization therapy with ventricular fusion pacing for electrical resynchronization in heart failure. Clinical Electrophysiology 7 (2021) 881–892.

