## Supplementary Materials for "Combination of personalized computational modeling and machine-learning for optimization of left ventricular pacing site in cardiac resynchronization therapy"

### Supplementary Material

#### CLINICAL DATA DESCRIPTION

In this study, we enrolled the clinical data from 57 CHF patients. All patients were on optimal drug treatment and underwent CRT device implantation at Almazov National Medical Research Centre between August 2016 and August 2019. Participants signed approved informed form. The study protocol was approved by the Institutional Ethical Committee.

The criteria for patient inclusion in the study were:

1. age over 18;
2. functional class (FC) II-IV of CHF according to the classification of the New York Heart Association (NYHA) at the outpatient stage of treatment;
3. LV EF $\leq 35\%$ (Simpson);
4. QRS duration (QRSd) more than 120 ms;
5. sinus rhythm, left bundle branch block (LBBB);
6. optimal drug therapy.

The exclusion criteria were:

1. acute myocardial infarction, transient ischemic attack, acute cerebrovascular accident less than 3 months before the start of the study;
2. patients who were scheduled to undergo myocardial revascularization or heart transplantation during the observation period;
3. congenital and acquired defects, as well as heart tumors, LV aneurysm, if scheduled for surgery during the observation period;
4. active inflammatory and autoimmune diseases of the myocardium;
5. thyrotoxicosis at the time of inclusion in the study;
6. anemic syndrome: blood hemoglobin level less than 90 g/l;
7. diseases limiting life expectancy to less than 1 year.

Patients underwent investigation according to standard pro forma with some additional research methods appropriate for this study.

The standard research methods included:

- clinical examination (complaints, medical history, physical examination) - before CRT and 1 year after CRT;
- general blood test, biochemical blood test (glucose, potassium, sodium, creatinine, urea, total bilirubin and its fractions, total cholesterol, total protein, AST, ALT), general urinalysis - before CRT;
- 12-lead ECG - before and 1 year after CRT; ECG monitoring during CRT device programming and during the entire observation period;
- echocardiographic studies before and 1 year after CRT to assess LV reverse remodeling;

- stress tests to exclude/confirm coronary artery disease: stress echocardiography, bicycle ergometry or treadmill test, where clinically indicated;
- coronary angiography, where clinically indicated.

The additional research methods included:

- ECG recording in intrinsic rhythm and under BiV pacing, while programming the CRT device within 7 days after implantation.
- Electrocardiographic imaging using an Amycard system (Amycard, EP Solutions SA, Yverdon, Switzerland). Prior to ECG imaging, a maximum of 224 unipolar body surface mapping electrodes were placed on the patient's torso, followed by computed tomography (CT) imaging of the heart and thorax (Somatom Definition 128, Siemens Healthcare, Germany). Subsequently, the electrodes were connected to the model 01C multichannel electrophysiology laboratory system (Amycard) for continuous ECG recordings during the pacing protocol. CT data were imported into Wave program version 2.14 (Amycard software) to reconstruct 3-dimensional geometry of the torso and heart. Finally, epi/endo ventricle models were manually built with marked active poles of RV and LV leads for bi-ventricular pacing simulations.
- MRI (MAGNETOM Trio A Tim 3 T, Siemens AG or INGENIA 1.5 T, Philips) with contrast (Gadovist or Magnevist) before CRT to detect structural damage of the myocardium.
- Tissue Doppler echocardiography to record ventricle mechanical dyssynchrony. Analysis of interventricular dyssynchrony (IVD) and intraventricular dyssynchrony in the LV (LVD) was performed using biomarkers suggested by Yu and co-authors **?**. IVD was assessed by the time difference between the start of systolic flows into the aorta and the pulmonary trunk as measured by a pulse-wave Doppler, a value of less than 40 ms was taken as an IVD normal value. LVD was assessed using two biomarkers: dyssynchrony index defined as the temporal difference between the maximal and minimal peak systolic velocities between 12 LV segments (Tsmax–Ts min, 105 ms was taken as threshold normal value), and standard deviation in the peak systolic velocities for 12 LV segments (SD–12, 34 ms was taken as cutoff value). To determine the peak systolic velocities, the technique of colour tissue Doppler ultrasonography was used.

Baseline clinical data for the patient cohort are presented in Table S1.

#### SUMMARY OF CLINICAL AND SIMULATION DATA IN THE PATIENT COHORT

**Table S1.** Clinical, imaging, model data and ML scores of CRT response in the patient cohort at baseline and BiV pacing with clinical lead configuration (ref-PS).

| Variable | All patients (n=57) | | | |
|---|---|---|---|---|
| | | **Clinical data** | | |
| Gender (male/female) | | 38/19 | | |
| Age, year | | 63±6 | | |
| BMI | | 28±5 | | |
| IHD/DCM | | 36 (63%)/21(37%) | | |
| History of AF | | 12 (21%) | | |
| | **LBBB** | **CRT** | **Δ, %** | **P*** |
| FC CHF : | | decrease in FC 31 (54%) | | |
| I | 0 (0%) | 10 (17.5%)* | 10 | **0.002** |
| II | 24 (42%) | 31 (54.5%) | 7 | 0.052 |
| III | 33 (58%) | 6 (10%)* | -27 | **0.001** |
| QRSd, ms | 195[175;204] | 157[147;175]* | -17[-29;-7] | **0.000** |
| | | **Echocardiography data** | | |
| EDV, ml | 278[235;341] | 223[161;270]* | -16[-39;-4] | **0.000** |
| ESV, ml | 202[157;257] | 136[101;177]* | -26[-51;-5] | **0.000** |
| EDD, mm | 73±8 | 66±10* | -10±10 | **0.000** |
| ESD, mm | 63±9 | 53±12* | -15±17 | **0.000** |
| EF, % | 26±6 | 35±8* | 9±8 | **0.000** |
| | | **CT/MRI data** | | |
| MTV,ml | | 338[240;412] | | |
| ScarV, ml | | 42[24;64] | | |
| ScarV/MTV | | 0.12[0.07;0.23] | | |
| Scar-LVPS distance, mm | | 34[10;49] | | |
| | | **Model data** | | |
| | **LBBB** | **BiV** | **Δ,%** | **P*** |
| TAT95, ms | 151[137;166] | 99[88;111]* | -37[-43;-26] | **0.000** |
| QRSd, ms | 190[175;205] | 142[132;155]* | -26[-32;-17] | **0.000** |
| $AD_{RVLV}$, ms | 72[49;82] | -3[-17;15]* | -106[-129;-84] | **0.000** |
| $AD_{STLV}$ | 0.26[0.22;0.29] | -0.08[-0.15;0.00]* | -133[-163;-100] | **0.000** |
| | | **Predictive model score** | | |
| ML-score | | 0.31[0.14;0.68] | | |

Mean±SD for normal distribution, median [ 25th percentile; 75th percentile] for non-normal distribution.
P* - LBBB *vs* CRT or LBBB *vs* BiV.
Comparisons between two dependent groups for quantitative data were made using Paired sample t-test for normal distribution and Wilcoxon's test for non-normal distribution. Comparisons between two dependent groups for qualitative data were made by McNemar's test.
Δ - Average change in indicator $\Delta X = X_{CRT} - X_{LBBB} / X_{LBBB}$ or $\Delta X = X_{BiV} - X_{LBBB} / X_{LBBB}$. The Δ is calculated as the absolute difference for normalized values (LV EF, $AD_{STLV}$), and for FC.
BMI – body mass index; IHD – ischemic heart disease; DCM – dilated cardiomyopathy; AF – atrial fibrillation; FC CHF – functional class of congestive heart failure; MTV – myocardial tissue volume; ScarV – scar/fibrosis volume; Scar-LVPS distance – distance between LV lead and scar/fibrosis area; TAT95 – 95% of total ventricular activation time; QRSd - maximal duration of QRS complex on 12 leads; $AD_{RVLV}$ - difference of total LV and RV activation time; $AD_{STLV}$ - difference between mean activation time of LV free wall and septum; ML-score - ML score of CRT response defined as more than 10% LV EF improvement.

**Table S2.** Clinical, imaging, model data and ML-scores of CRT response in responder and non-responder subgroups. CRT response is defined as more than 10% improvement in the LV EF under BiV pacing with clinical (referent) lead position (ref-PS) in a year after CRT device implantation.

| | Patient cohort n=57 | | | | | |
|---|---|---|---|---|---|---|
| Variable | **Responders n=23 (40%)** | | | **Non-responders n=34 (60%)** | | |
| | | | **Clinical data** | | | |
| Gender (male/female) | 15/8 | | | 23/11 | | |
| Age, year | 64±6 | | | 63±7 | | |
| BMI | 27±5 | | | 30±5# | | |
| IHD/DCM | 14 (61%)/9(49%) | | | 22 (65%)/12(35%) | | |
| History of AF | 4 (17%) | | | 8 (24%) | | |
| | **LBBB** | **CRT** | **Δ, %** | **LBBB** | **CRT** | **Δ, %** |
| FC CHF : | decrease in FC 17 (70%) | | | decrease in FC 15 (44%) | | |
| I | 0 (0%) | 7 (30%)* | 7 | 0 (0%) | 3 (9%) | 3 |
| II | 12 (52%) | 12 (52%) | 0 | 12 (35%) | 19 (56%)** | 7 |
| III | 11(48%) | 2 (8%)* | -9 | 22 (65%) | 4 (12%)** | -18 |
| QRSd, ms | 192±20 | 143±14** | -19[-29;-8] | 190±26 | 145±21** | -15[-28;-6] |
| | | | **Echocardiography data** | | | |
| EDV, ml | 314[252;355] | 178[134;241]** | -29[-50;-14] | 259[223;330] | 231[173;286]## | -9[-23;7]## |
| ESV, ml | 232[196;267] | 118[78;142]** | -46[-61;-31] | 180[151;247] | 152[113;199]*## | -9[-25;5]## |
| EDD, mm | 74±8 | 62±10** | -16±10 | 73±7 | 69±9 **## | -5±8 ## |
| ESD, mm | 64±9 | 48±13** | -26±17 | 62±9 | 57±10 **## | -7±13## |
| EF, % | 23±5 | 40±6 ** | 16[14;19] | 29±6## | 32±7**## | 4[1;8]## |
| | | | **CT/MRI data** | | | |
| MTV,ml | 294[229;396] | | | 361[272;469] | | |
| ScarV, ml | 34[25;50] | | | 48[21;84] | | |
| ScarV/MTV | 0.13[0.08;0.18] | | | 0.11[0.06;0.29] | | |
| Scar-LVPS distance, mm | 44[19;53] | | | 25[3;43]# | | |
| | | | **Model data** | | | |
| | **LBBB** | **BiV** | **Δ, %** | **LBBB** | **BiV** | **Δ,%** |
| TAT95, ms | 153[141;164] | 96[88;106]** | -37[-41;-28] | 150[133;169] | 102[88;112]** | -37[-43;-19] |
| QRSd, ms | 195[180;204] | 142[132;151]** | -26[-30;-22] | 189[173;210] | 147[131;156]** | -26[-33;-15] |
| $AD_{RVLV}$, ms | 70[52;84] | -8[-16;7]** | -117[-133;-88] | 73[47;83] | 2[-18;17]** | -98[-127;-82] |
| $AD_{STLV}$ | 0.24[0.20;0.27] | -0.09[-0.15;-0.03] | -143[-165;-110] | 0.26[0.22;0.29] | -0.08[-0.14;0.01]** | -129[-160;-97] |
| | | | **Predictive model score** | | | |
| ML-score | 0.73[0.36;0.95] | | | 0.19[0.07;0.39]## | | |

Mean±SD for normal distribution, median [ 25th percentile; 75th percentile] for non-normal distribution.
* - p<0.05, ** - p<0.01 LBBB *vs* CRT or LBBB *vs* BiV. Comparisons between two dependent groups were made using Wilcoxon's test for quantitative data and McNemar's test for qualitative data.
### - p<0.05, ## - p<0.01 Responders *vs* Non-responders. Comparison between two independent groups was carried out using the Mann-Whitney test for quantitative data and Pearson's chi-square test for qualitative data.

Δ - Average change in indicator ΔX = X CRT - X LBBB / XLBBB or ΔX = XBiV - X LBBB / X LBBB. Δ is calculated as the absolute difference for normalized values (EF and $AD_{STLV}$) and FC
BMI - Body mass index; IHD -Ischemic heart disease; DCM - Dilated cardiomyopathy; AF - Atrial Fibrillation; FC CHF- functional class of congestive heart failure; MTV - myocardial tissue volume; ScarV – scar/fibrosis volume; Scar-LVPS distance – distance between LV lead and scar/fibrosis area; TAT95 - 95% of total ventricular activation time; QRSd – maximal QRS duration in 12-lead ECG; $AD_{RVLV}$ – difference of total LV and RV activation time; $AD_{STLV}$ – difference between mean activation time of LV free wall and septum; ML-score – ML score of CRT response defined as more than 10% LV EF improvement.

#### LV PACING SITE LOCATION

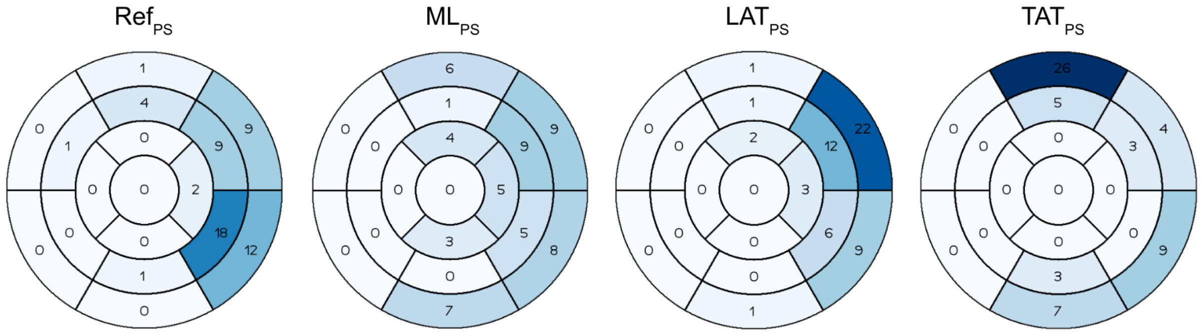


**Figure S1.** Distribution of the LV pacing site locations in the 17–segment AHA model for different pacing designs in the patient cohort. The number of patients (out of 57) with pacing lead located in the segment is indicated.

#### SCARRING AREA IN THE LEFT VENTRICLE

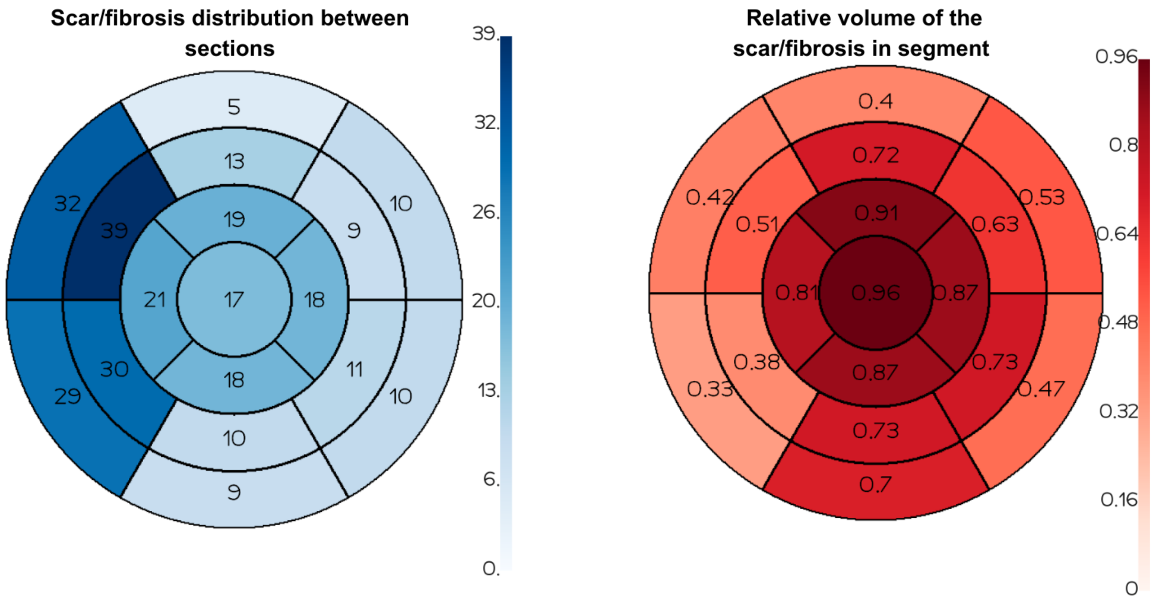


**Figure S2.** Distribution of the simulated scar/fibrosis area between 17 segments in AHA LV model based on MRI data in the patient cohort. Left panel shows the number of patients (out of 57 total) with scar/fibrosis area in each segment of 17-segment AHA LV models. Right panel shows a relative volume of damaged myocardium in the segments.

#### ML CLASSIFIER OF CRT RESPONSE

Figure S3 shows a list of clinic and simulated features used to develop logistic regression classifier (LR) in the range of their significance for CRT response prediction (right panel). A ROC curve for the final LR classifier based on selected 7 most significant features (see red lines on the right) with ROC AUC of 0.84.

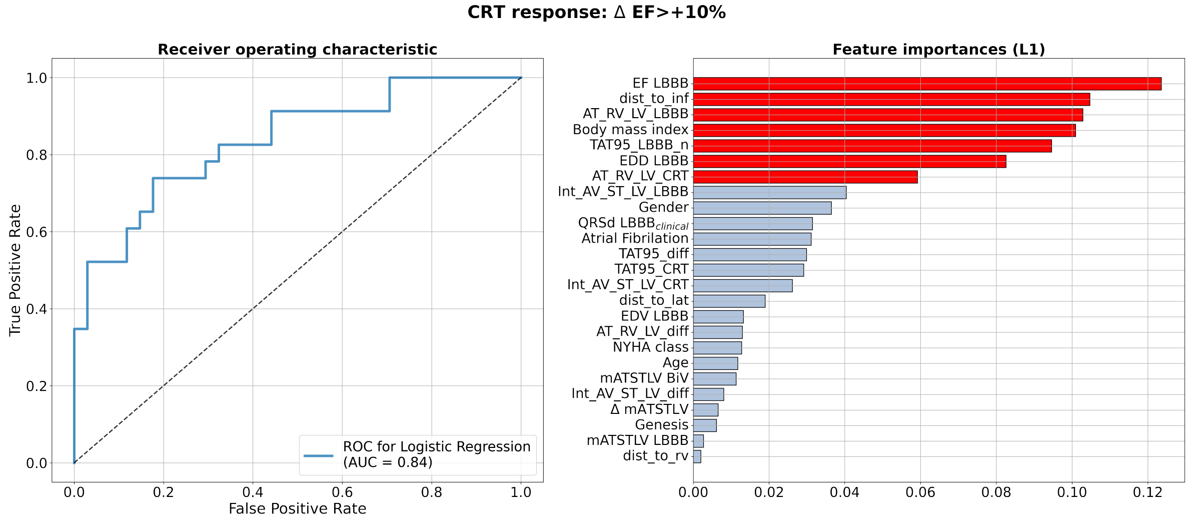


**Figure S3.** Machine Learning Classifier for CRT response prediction from the hybrid dataset of clinical and model-derived data for 57 patients. **Left panel** shows receiver operating characteristic (ROC) curves for the ML classifier based on the $\Delta$EF>10% criterion of CRT response. Value of the area under the ROC curve (ROC AUC) for the model is shown on the panel. **Right panel** lists clinical and model-derived features in the descending order of importance ranged using a logistic regression feature selection approach for the ML classifier.

A list of features and corresponding logistic regression (LR) weights in the final LR classifier are shown in TableS3. The LR classifier generates an ML-score based on the clinical and simulated features of the patients. Cross-table and characteristics of the LR classifier are shown below.

**Table S3.** Logistic Regression classifier of CRT response defined as more than 10% improvement in LV EF

| Feature | Data type | Non-standardized coefficient of LR | Standardized coefficient of LR |
|---|---|---|---|
| EF, % | Clinical | -15.33 | -1.16 |
| $AD_{RVLV}$ LBBB, ms | Simulated | -0.04 | -0.98 |
| BMI | Clinical | -0.19 | -0.90 |
| Scar-LV distance, mm | Simulated | 0.03 | 0.84 |
| TAT_LBBB/MTV | Simulated | 3.96 | 0.75 |
| EDD, mm | Clinical | 0.08 | 0.70 |
| $AD_{RVLV}$ BiV, ms | Simulated | -0.02 | -0.37 |
| Intercept of LR | | 2.76 | -0.51 |

LR - Logistic Regression
BMI - Body mass index; TAT95 - 95% of total ventricular activation time; MTV - myocardial tissue volume; $AD_{RVLV}$ - difference of total LV and RV activation time; EDD – end diastolic diameter of LV ; EF – ejection fraction.

**Table S4.** LR classifier performance

| ROC AUC =0.84, cut-off = 0.5 | | Clinic Responder | Clinic Non-responder | |
|---|---|---|---|---|
| LR positive (ML-SCR >0,5) | | 15 | 5 | |
| LR negative (ML-SCR <0,5) | | 8 | 29 | |
| Accuracy | Sensitivity | Specificity | ppv | npv |
| 77% | 65% | 85% | 75% | 78% |

#### COMPARISON OF SIMULATED FEATURES IN DIFFERENT VENTRICULAR ACTIVATION DESIGNS

**Table S5.** Comparison of inter-ventricular ($AD_{RVLV}$) and intra-ventricular ($AD_{STLV}$) electrical dyssynchrony indices at baseline LBBB and BiV pacing with different LV lead positions in the total patient cohort and subgroups of responders and non-responders

| Simulated feature | | Value | Difference (BiV – LBBB) | Difference (ML-PS – ref-PS) |
|---|---|---|---|---|
| | | **Total Cohort (n=57)** | | |
| $AD_{RVLV}$, ms | LBBB | 72[49;82] | | |
| | ref-PS | -3[-17;15]** | -69[-89;-53] | |
| | ML-PS | 4[1;12]** | -61[-78;-43]$ | 9[-2;21] |
| $AD_{STLV}$ | LBBB | 0.26[0.22;0.29] | | |
| | ref-PS | -0.08[-0.15;0.00]** | -0.30[-0.40;-0.23] | |
| | ML-PS | 0.07[0.04;0.15]**$$ | -0.18[-0.21;-0.11]$$ | 0.15[0.07;0.23] |
| | | **Responders (n=23)** | | |
| $AD_{RVLV}$, ms | LBBB | 70[52;84] | | |
| | ref-PS | -8[-16;7]** | -73[-88;-50] | |
| | ML-PS | 6[2;11]**$ | -61[-74;-42]$ | 10[0;21] |
| $AD_{STLV}$ | LBBB | 0.24[0.20;0.27] | | |
| | ref-PS | -0.09[-0.15;-0.03]** | -0.30[-0.38;-0.24] | |
| | ML-PS | 0.08[0.02;0.18]**$$ | -0.15[-0.22;-0.07]$$ | 0.14[0.08;0.28] |
| | | **Non-responders (n=34)** | | |
| $AD_{RVLV}$, ms | LBBB | 73[47;83] | | |
| | ref-PS | 2[-18;17]** | -67[-90;-54] | |
| | ML-PS | 3[1;12]** | -63[-82;-42]$$ | 3[-4;19] |
| $AD_{STLV}$ | LBBB | 0.26[0.22;0.29] | | |
| | ref-PS | -0.08[-0.14;0.01]** | -0.31[-0.42;-0.23] | |
| | ML-PS | 0.07[0.05;0.12]**$$ | -0.18[-0.21;-0.14] | 0.16[0.05;0.22] |

Median [ 25th percentile; 75th percentile]
* - p<0.05, ** - p<0.01 ML-PS *vs* LBBB. $ - p<0.05, $ $ - p<0.01 ML-PS vs ref-PS . & - p<0.05, && - p<0.01 ML-PS vs ML-PS . Comparison of dependent groups was performed using Friedman's test, followed by a pairwise comparison adjusted for multiple comparisons.
### - p<0.05, # - p<0.01 Responders vs Non-responders. Comparison between two independent groups was carried out using Mann-Whitney test. $AD_{RVLV}$ - difference of total LV and RV activation time; $AD_{STLV}$ - difference between mean activation time of LV free wall and septum.

**Table S6.** Comparison of model-derived features at different LV pacing lead positions in the total patient cohort and subgroups of responders and non-responders

| Simulated feature | | Total Cohort (n=57) | | Responders (n=23) | | Non-responders (n=34) | |
|---|---|---|---|---|---|---|---|
| | | Value | Difference (ML-PS – ref-PS) | Value | Difference (ML-PS – ref-PS) | Value | Difference (ML-PS – ref-PS) |
| RV-LV delay,ms | ref-PS | 110[75;126] | | 112[96;130] | | 98[66;124] | |
| | ML-PS | 89[68;136] | -4[-31;13] | 84[70;136] | -8[-39;14] | 93[65;134] | -3[-30;14] |
| RV-LV distance, mm | ref-PS | 102[85;115] | | 102[84;113] | | 97[87;119] | |
| | ML-PS | 97[80;120] | 0[-19;15] | 101[76;109] | -5[-24;15] | 96[82;126] | 2[-9;15] |
| LAT-LVPS distance,mm | ref-PS | 58[34;70] | | 59[33;66] | | 55[34;70] | |
| | ML-PS | 76[57;95]$ | 17[1;46] | 82[71;93]$ | 17[5;49] | 74[51;101] | 15[-5;45] |
| Scar-LVPS distance,mm | ref-PS | 34[10;49] | | 44[19;53] | | 25[3;43]# | |
| | ML-PS | 58[45;71]$$ | 23[9;35] | 61[51;66]$ | 17[8;33] | 55[41;72]$$ | 27[12;46] |

Median [ 25th percentile; 75th percentile]
$ - p<0.05, $$ - p<0.01 ML-PS vs ref-PS . & - p<0.05 - ML-PS vs ML-PS . Comparison of dependent groups was performed using Friedman's test, followed by a pairwise comparison adjusted for multiple comparisons.
### - p<0.05 Responders vs Non-responders. Comparison between two independent groups was carried out using Mann-Whitney test.

#### DISTRIBUTION OF CLINICAL RESPONDERS AND NON-RESPONDERS BETWEEN SUBGROUPS CLASSIFIED AS POSITIVE OR NEGATIVE TO CRT RESPONSE

In Table S7 we show a cross-table of LR classifier predictions of positive (ML-score>0.5) and negative (ML-score <0.5) response to CRT for clinic responders and non-responders. Different colors show true and false positive (red and orange) and true and false negative (black and blue) in the patient cohort as classified from LR fed by simulations for the referent pacing lead positions (ref-PS). In the lower rows we show similar cross-tables for different optimal lead positions with indication of transitions between the subgroups.

The total ratio of patients classified as positive and negative to CRT response for each optimized LV pacing lead position is shown in the right column. It can be that only the ML-score based optimal LV lead position (ML-PS) increases the amount of positive to CRT response as compared to that observed in clinic.

**Table S7.** Cross-table between clinical responders/ non-responders and positive/negative prediction of CRT response at BiV pacing with different LV pacing lead positions

| PS | ML-score | Clinic | | |
|---|---|---|---|---|
| | | Responder 23 (40%) | Non-responder 34(60%) | Total |
| ref-PS | ML-SCR>0.5 (positive) | **15** | **5** | 20 (35%) |
| | ML-SCR<0.5 (negative) | **8** | **29** | 37 (65%) |
| ML-PS | ML-SCR>0.5 (positive) | **15**+**5**=20 | **5**+**6**=11 | 31 (54%) |
| | ML-SCR<0.5 (negative) | **8**-**5**=3 | **29**-**6**=23 | 26 (46%) |

ML-SCR - ML-score
**True Positive**
**False Negative by ref-PS**
**False Positive by ref-PS**
**True Negative**

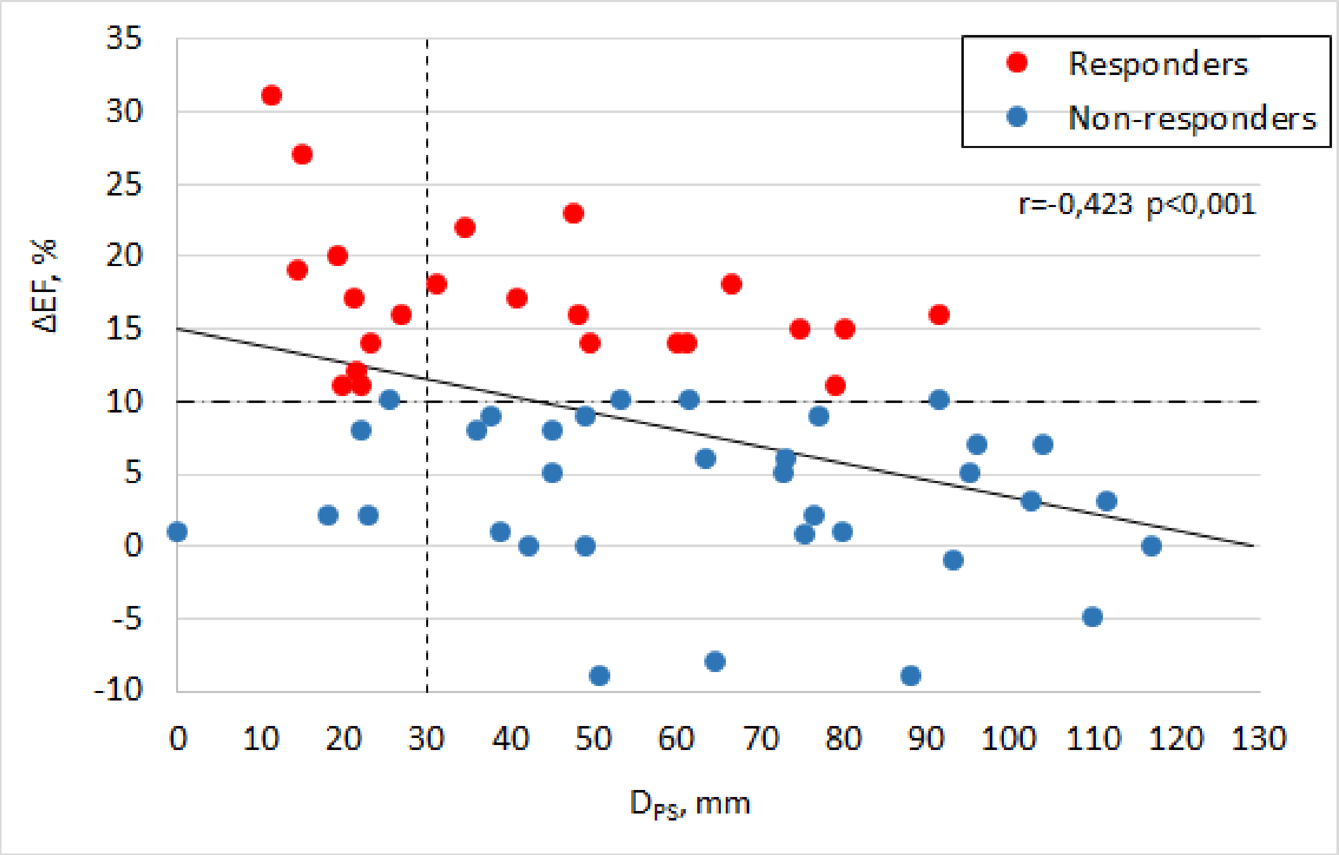


**Figure S4.** Regression between the change in EF and $D_{PS}$. r - Spearman's correlation coefficient, p - the coefficient significance. The dot dash line is the cutoff 10% for the ejection fraction (EF). The dotted line is the cutoff 30mm for the $D_{PS}$.The solid line is a linear regression trend.

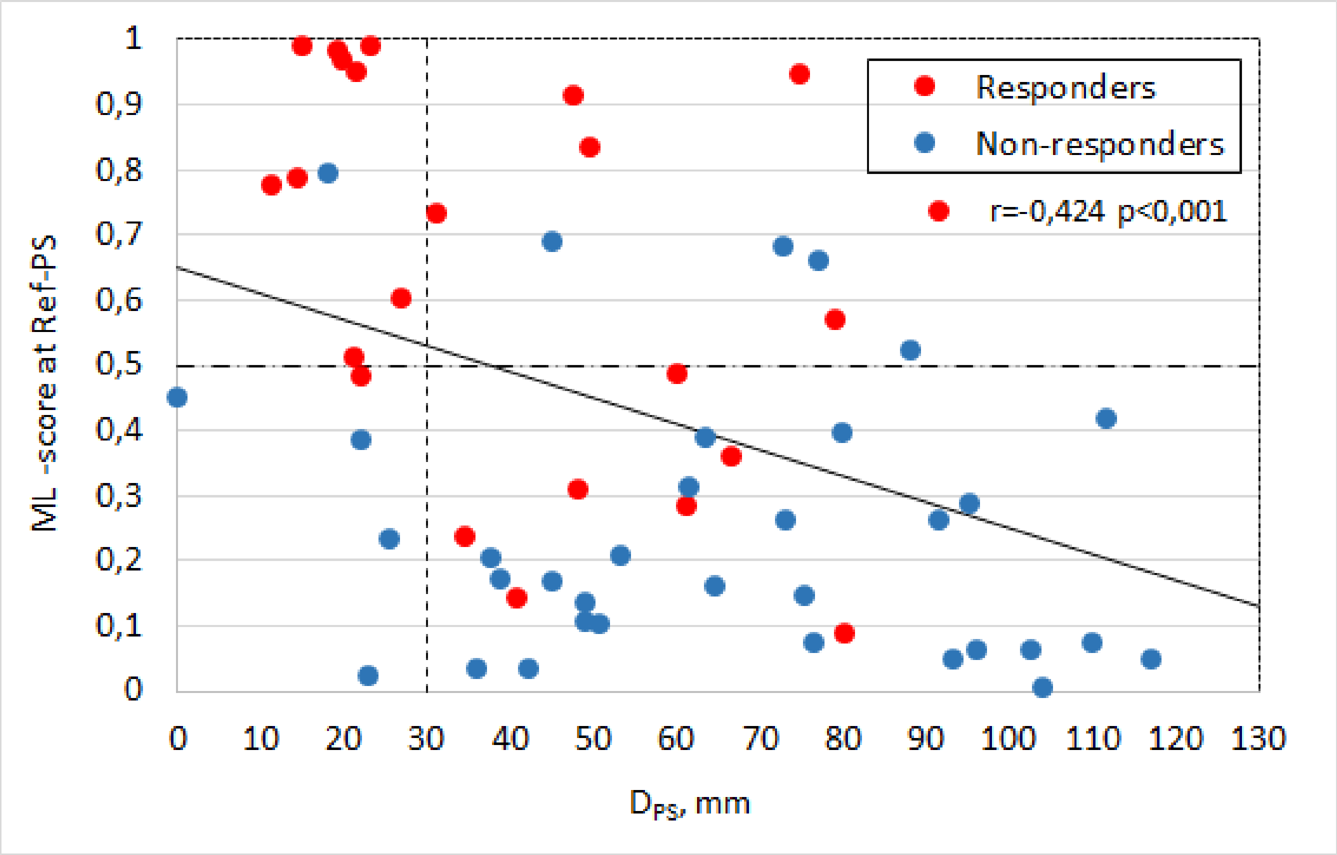


**Figure S5.** Regression between ML-score at Ref-PS and $D_{PS}$. r - Spearman's correlation coefficient, p - the coefficient significance. The dot dash line is the ML-score cutoff 0,5 . The dotted line is the cutoff 30mm for the $D_{PS}$.The solid line is a linear regression trend.

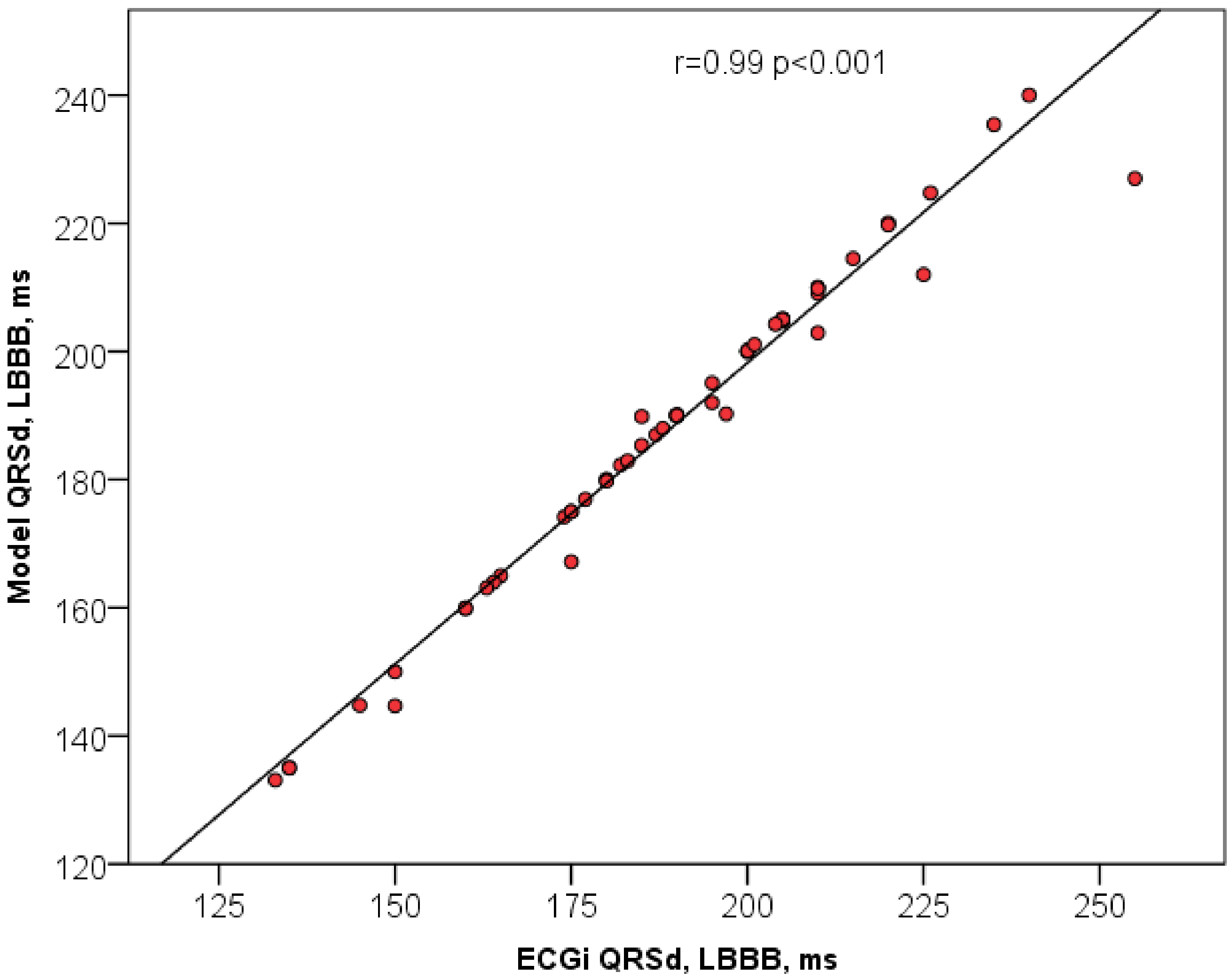


**Figure S6.** Regression between ECGi QRSd and Model QRSd at LBBB. r - Pearson's correlation coefficient, p - the coefficient significance. The solid line is a linear regression trend.

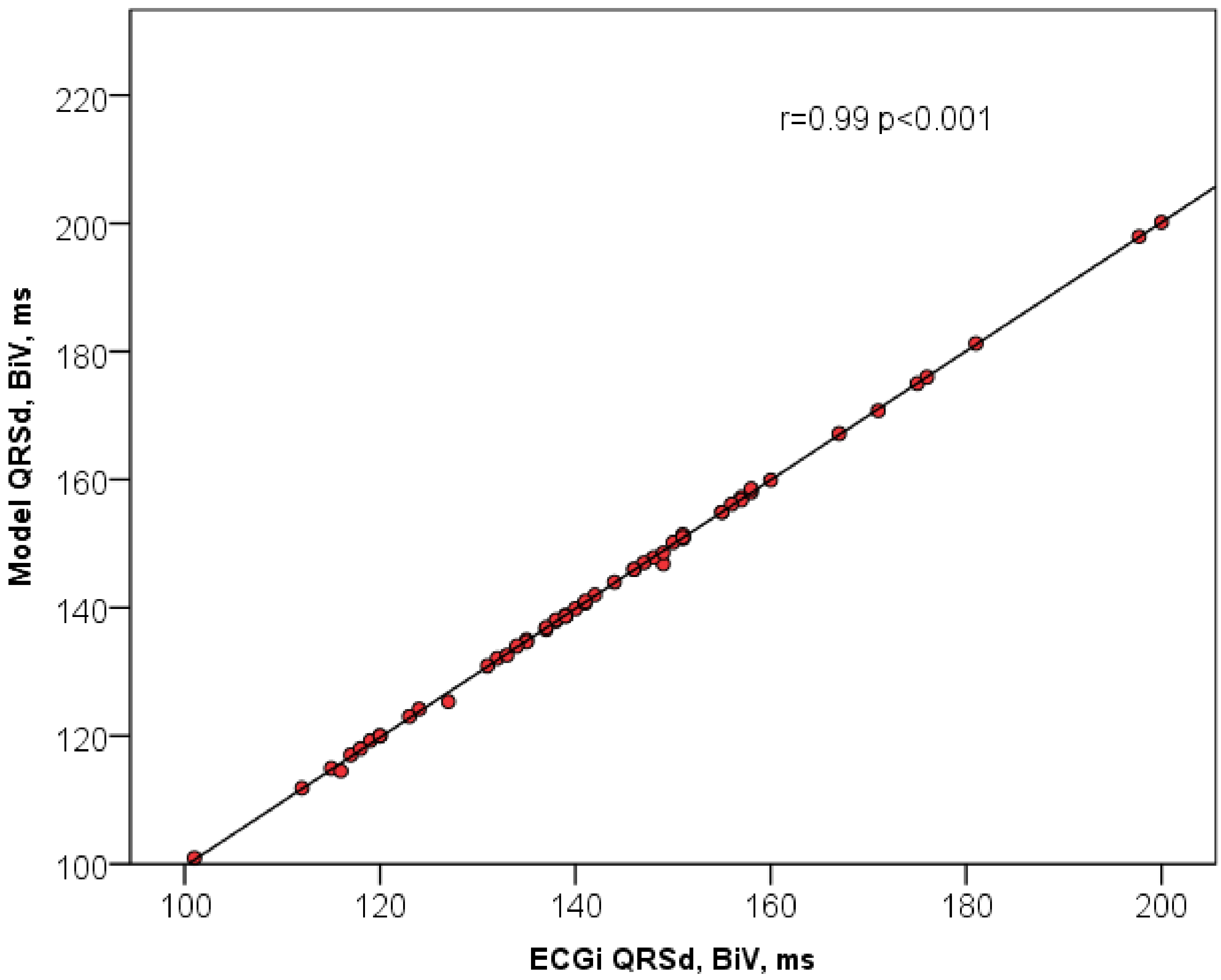


**Figure S7.** Regression between ECGi QRSd and Model QRSd at BiV. r - Pearson's correlation coefficient, p - the coefficient significance. The solid line is a linear regression trend.

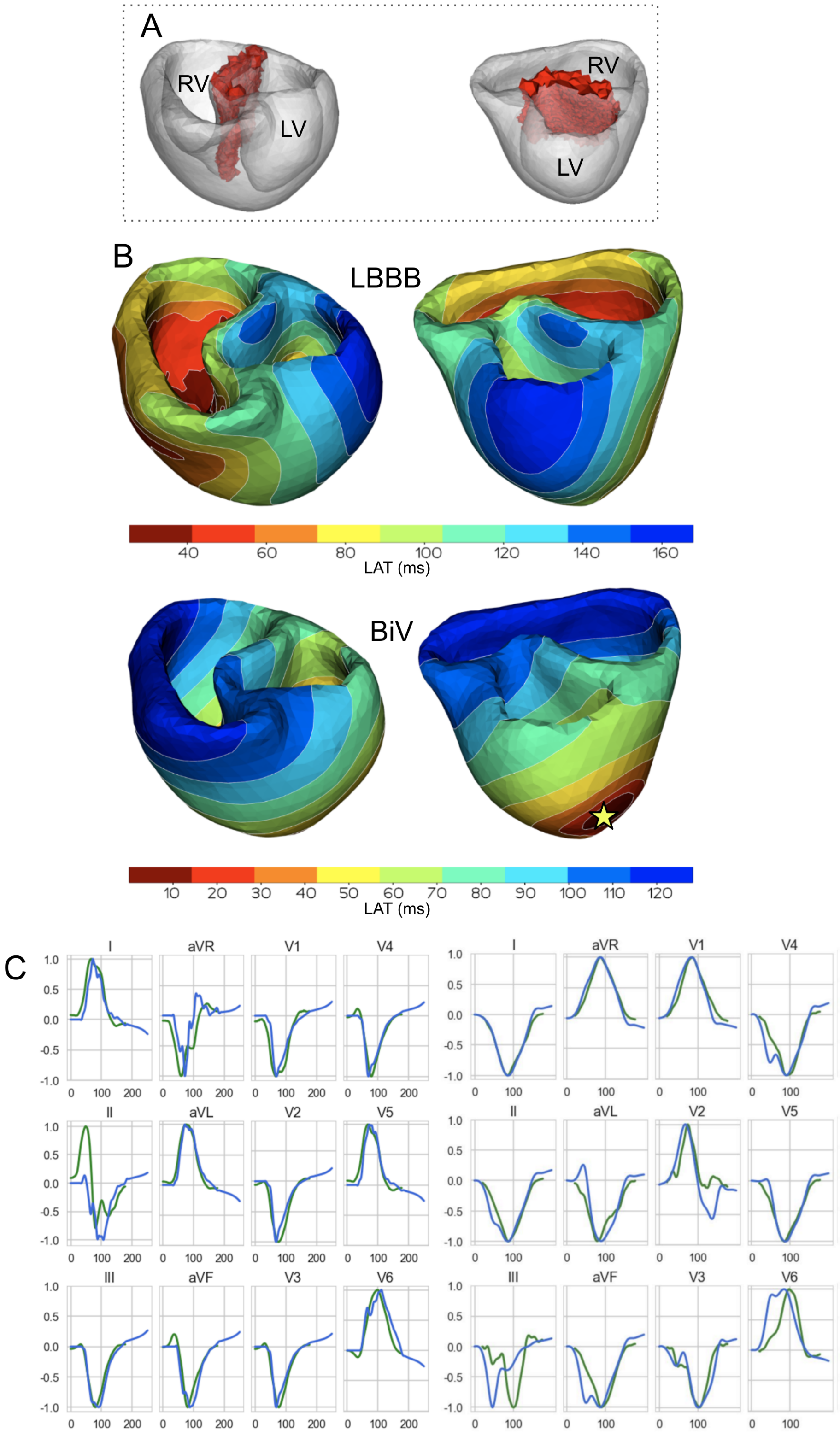


**Figure S8.** Model validation. Example of a personalized ventricular model for patient #11. A. Area of fibrosis in the interventricular septum (red zone). B. Comparison of model activation maps for LBBB (top) and BiV pacing (bottom). Star indicates LV pacing site for BiV pacing stimulation. The RV stimulating electrode was located in the apex of the surface. C. Calculated ECG signals (QRS complexes) for LBBB on the left and BiV pacing on the right. Green line - signals recorded in the clinic. Blue line - simulated signals. The amplitude of the QRS signals is normalized to the maximum values of the signals.

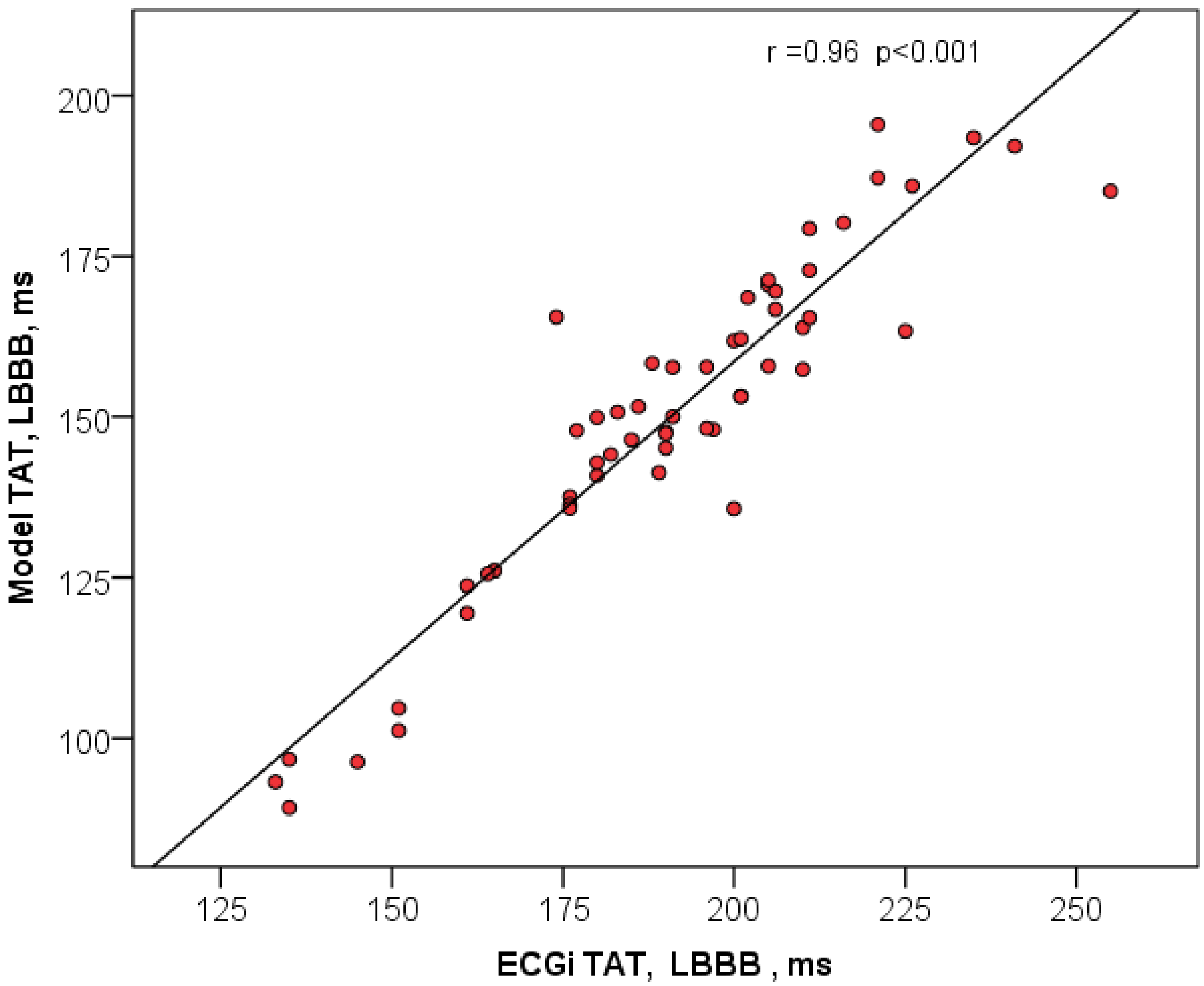


**Figure S9.** Regression between ECGi TAT and Model TAT at LBBB. r - Pearson's correlation coefficient, p - the coefficient significance. The solid line is a linear regression trend.

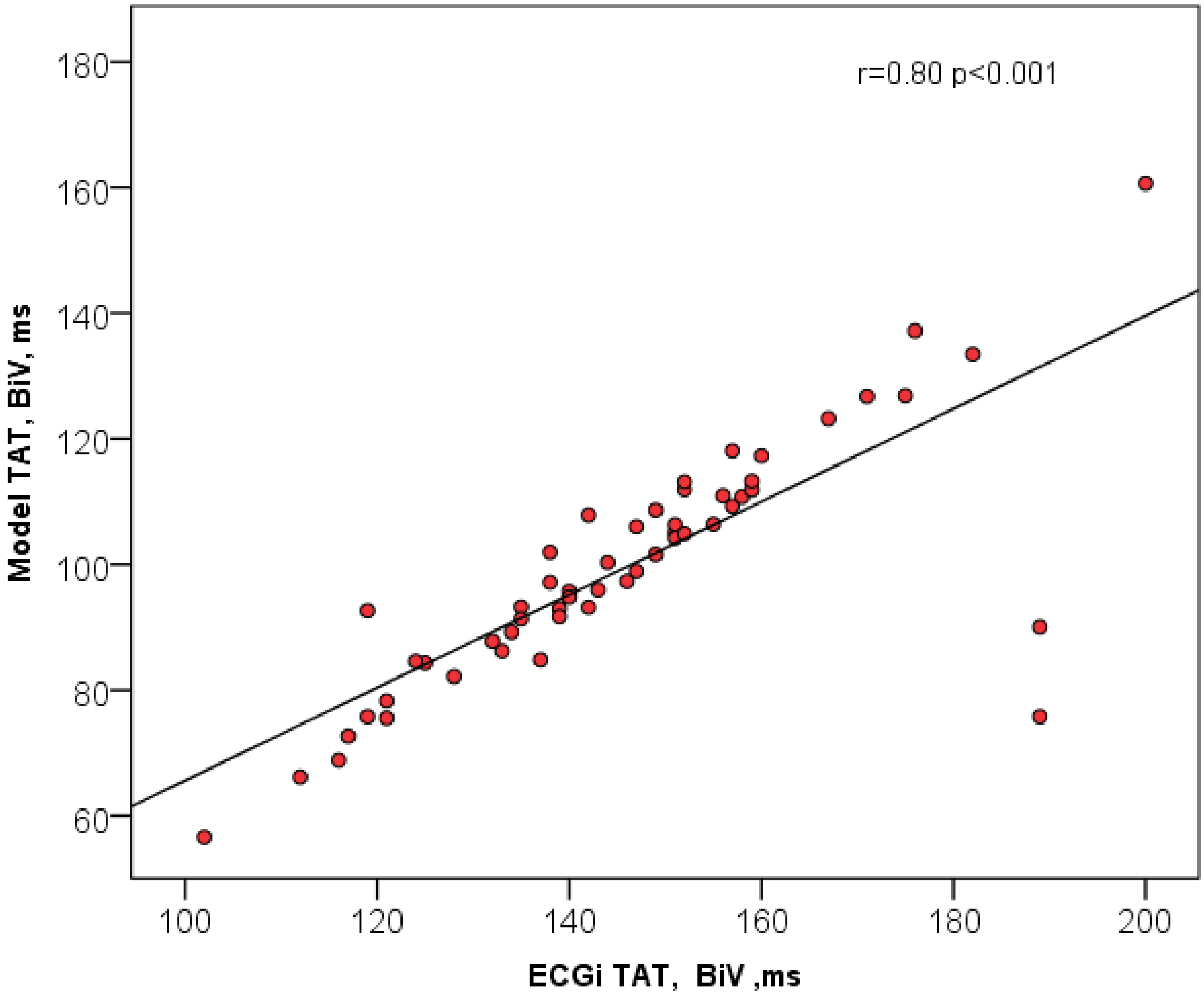


**Figure S10.** Regression between ECGi TAT and Model TAT at BiV. r - Pearson's correlation coefficient, p - the coefficient significance. The solid line is a linear regression trend.
